# Whole-Cerebrum distortion-free three-dimensional pseudo-Continuous Arterial Spin Labeling at 7T

**DOI:** 10.1101/2023.04.24.23289051

**Authors:** Chenyang Zhao, Xingfeng Shao, Qinyang Shou, Samantha J. Ma, Sayim Gokyar, Christina Graf, Rudolf Stollberger, Danny JJ Wang

## Abstract

Fulfilling potentials of ultrahigh field for pseudo-Continuous Arterial Spin Labeling (pCASL) has been hampered by B1/B0 inhomogeneities that affect pCASL labeling, background suppression (BS), and the readout sequence. This study aimed to present a whole-cerebrum distortion-free three-dimensional (3D) pCASL sequence at 7T by optimizing pCASL labeling parameters, BS pulses, and an accelerated Turbo-FLASH (TFL) readout. A new set of pCASL labeling parameters (Gave=0.4mT/m, Gratio=14.67) was proposed to avoid interferences in bottom slices while achieving robust labeling efficiency (LE). An OPTIM BS pulse was designed based on the range of B1/B0 inhomogeneities at 7T. A 3D TFL readout with 2D-CAIPIRINHA undersampling (R=2×2) and centric ordering was developed, and the number of segments (Nseg) and flip angle (FA) were varied in simulation to achieve the optimal trade-off between SNR and spatial blurring. In-vivo experiments were performed on 19 subjects. The results showed that the new set of labeling parameters effectively achieved whole-cerebrum coverage by eliminating interferences in bottom slices while maintaining a high LE. The OPTIM BS pulse achieved 33.3% higher perfusion signal in gray matter (GM) than the original BS pulse with a cost of 4.8-fold SAR. Incorporating a moderate FA (8^°^) and Nseg (2), whole-cerebrum 3D TFL-pCASL imaging was achieved with a 2×2×4 mm^3^ resolution without distortion and susceptibility artifacts compared to 3D GRASE-pCASL. In addition, 3D TFL-pCASL showed a good to excellent test-retest repeatability and potential of higher resolution (2 mm isotropic). The proposed technique also significantly improved SNR when compared to the same sequence at 3T and simultaneous multislice TFL-pCASL at 7T. By combining a new set of labeling parameters, OPTIM BS pulse, and accelerated 3D TFL readout, we achieved high resolution pCASL at 7T with whole-cerebrum coverage, detailed perfusion and anatomical information without distortion, and sufficient SNR.

## 1. Introduction

Arterial Spin Labeling (ASL) is a non-invasive MR imaging technique permitting the quantitative measurement of cerebral blood flow (CBF) (Detre et al., 1992; Williams et al., 1992). After thirty years of research, the community has reached a consensus for the clinical implementation at 3T recommending pseudo-Continuous ASL (pCASL), background suppression (BS), and segmented three-dimensional (3D) readout (Alsop et al., 2015). As a technique inherently suffering from low signal-to-noise ratio (SNR) and limited tracer half-life, ultrahigh-field (7T) is appealing to ASL given the potential advantages of increased SNR and prolonged blood T1. At 7T, ASL has been explored by several studies with different labeling and imaging techniques, such as 2D turbo-FLASH (TFL)-pCASL (Zuo et al., 2013), 2D TFL-pCASL with simultaneous multi-slice acquisition (SMS-TFL-pCASL) (Wang et al., 2015), 2D echo-planar imaging (EPI)-pCASL (Luh et al., 2013), pulsed ASL (Wang et al., 2021), and laminar perfusion imaging with a 3D zoomed gradient and spin echo (GRASE) sequence (Shao et al., 2021). Nevertheless, 3D pCASL imaging with whole-brain coverage and high resolution remains difficult at 7T because of the following challenges.

The labeling efficiency (LE) of pCASL is low due to the limited coverage of transmit coil. To address this issue, placing the labeling plane at a more superior position at 7T than 3T was recommended (Zuo et al., 2013). A recent study also recommended a smaller average gradient (G_ave_) and slice-selective gradient (G_max_) of pCASL labeling to compensate for a potentially reduced B1 field at the labeling region (Meixner et al., 2022). In addition, an increased duty cycle of pCASL labeling was suggested to improve the robustness of LE to B0 inhomogeneity (Wang et al., 2022). However, the combination of these modifications may produce signal interferences in bottom imaging slices because of sidebands of pCASL labeling extending into the imaging volume, which limits field-of-view (FOV) in the slice direction.

In addition, the utility of BS is limited by the low inversion efficiency (IE) due to B1/B0 field inhomogeneities at 7T. BS is crucial for improving temporal SNR (tSNR) and suppressing artifacts related to physiological fluctuations. At 3T, the reduction of ASL signal caused by a BS pulse is negligible (<5%) (Garcia et al., 2005), and multiple BS pulses can be applied to heavily suppress different tissues (Shao et al., 2018). At 7T, however, the use of BS, in particular multiple BS pulses, may lead to a substantial loss of ASL signal. On the other hand, BS is important for segmented 3D ASL at 7T as physiological noise becomes more dominant than at 3T. Consequently, a BS pulse with high IE and resistance to B1/B0 field inhomogeneities is highly desired.

Furthermore, workhorse readout sequences for pCASL at 3T, such as GRASE and EPI (Alsop et al., 2015), suffer from signal loss and distortion due to shortened T2/T2* at 7T. A previous study proposed GRASE-pCASL using 12-fold acceleration to reduce TE to ∼20 ms and total-generalized-variation (TGV) regularized reconstruction to improve SNR (Shao et al., 2020; Spann et al., 2020). Alternatively, TFL is a readout sequence with successful applications in structural (Marques et al., 2010) and susceptibility weighted (Bian et al., 2016) MRI at 7T due to the advantages of short TE, fast acquisition time, no or minimal distortion, and low specific absorption rate (SAR) of RF power. Comparing to the previous studies that used 2D and 2D SMS-TFL-pCASL (Wang et al., 2022, 2015; Zuo et al., 2013), 3D TFL is preferred for achieving higher resolution and SNR.

This work aimed to develop a distortion-free 3D pCASL sequence with whole-cerebrum coverage by combining a new set of pCASL labeling parameters, a high-performance BS pulse, and an accelerated TFL readout at 7T. Firstly, theoretical analysis and Bloch equation simulation were performed to propose an optimal set of pCASL parameters with high LE and without interferences with bottom imaging slices. Secondly, Bloch simulations were conducted to evaluate several BS pulses from which a BS pulse with superior performance was selected.

Thirdly, TFL-pCASL was compared with GRASE-pCASL based on simulations, which were validated by in-vivo experiments. Moreover, the test-retest repeatability was examined. Finally, in-vivo experiments were conducted to compare the proposed technique to the same sequence at 3T and SMS-TFL-pCASL at 7T.

## 2. Method

### 2.1. Optimization of pCASL labeling

According to (Dai et al., 2008), the pCASL pulse train is formed as the convolution of Hanning function, ℎ(𝑡), and a temporal sampling function, 𝑥(𝑡). Its Fourier response, 𝐹(𝑧), can be derived as the multiplication of 𝐻(𝑧) and X(𝑧), which are Fourier responses of ℎ(𝑡) and 𝑥(𝑡), respectively. The distance from the labeling plane, denoted by z, is associated with frequency along the z gradient. Subscripts of “con” and “la” were used to indicate the control and label conditions, respectively, for expressions throughout the paper. The signs of 𝑋_*con*_(𝑡) alternate between positive and negative, resulting in a X_*con*_(𝑧) which is shifted half cycles from X_$%_(𝑧). Expressions of the functions are described in supplement section 1.

Figure 1 shows the aforementioned functions with the benchmark parameter set (balanced scheme, RF spacing (T)=550 µs, RF duration (𝜏)=300 µs, G_ave_=0.6 mT/m, and G_max_/G_ave_ (G_ratio_)=10), herein termed Labeling 0, that was previously optimized in (Wang et al., 2022). Spikes in 𝐹_*con*_(𝑧) appear at the off-center locations where frequency peaks of X_*con*_(𝑧) overlap with the sidelobes of 𝐻(𝑧). Similarly, two smaller spikes in 𝐹_$%_(𝑧) appear at different locations. In the original implementation of pCASL at 3T, the harmonic frequency peaks of X(𝑧) appeared at distances far enough, where negligible sidelobes of 𝐻(𝑧) reside, to avoid the formation of spikes in 𝐹(𝑧) within the imaging FOV. At 7T, however, the modifications for optimizing LE mentioned in the introduction may induce spikes in 𝐹(𝑧) within FOV, thus interfering perfusion signal in 𝐹_*con*_(𝑧) − 𝐹_$%_(𝑧). Since X_*con*_(𝑧) generates ∼10 times stronger interferences than X_$%_(𝑧), the following analysis will focus on the control condition.

**Figure 1.**
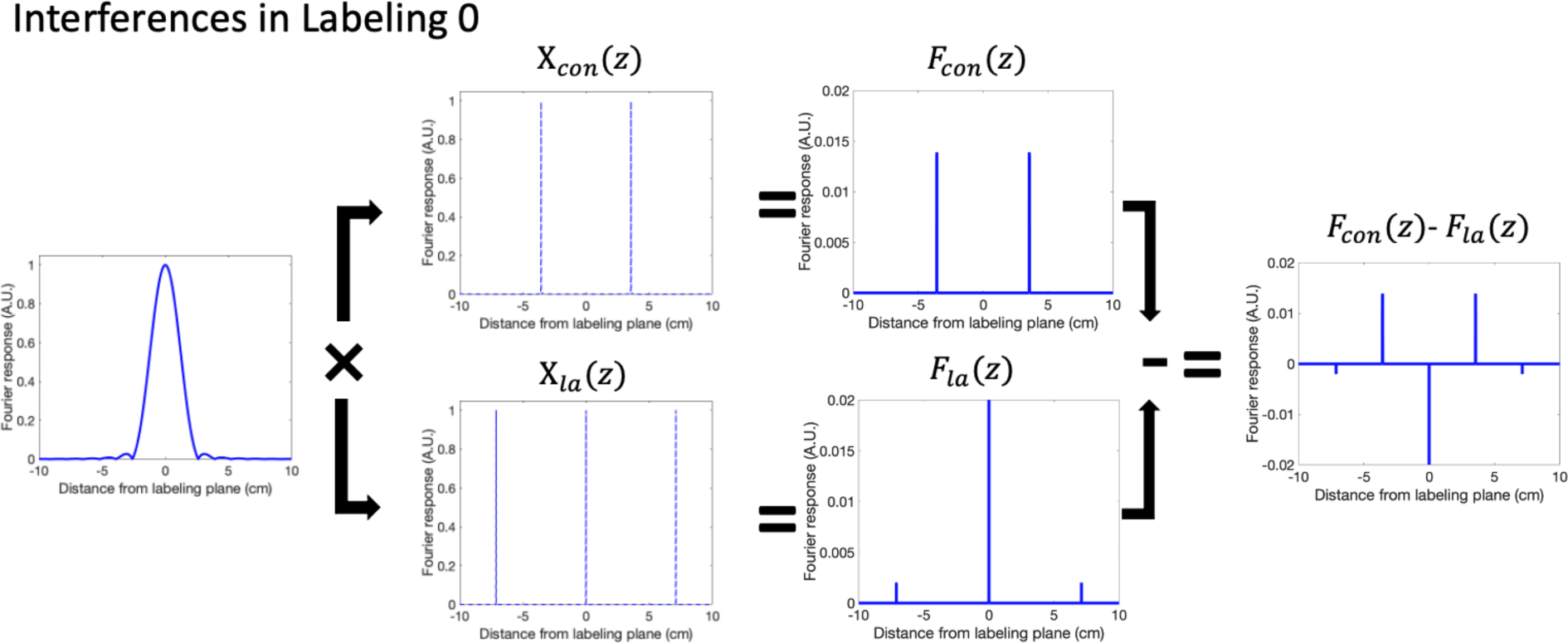
A schematic plot showing the formation of signal interferences in Labeling 0. 𝐻(𝑧) and 𝑋(𝑧) are Fourier responses of Hanning function, ℎ(𝑡), and a temporal sampling function, 𝑥(𝑡), respectively. Harmonic frequency peaks of 𝑋(𝑧) overlap with the sidelobes of 𝐻(𝑧), which generates spikes in 𝐹(𝑧) on static tissues. The differences of 𝐹(𝑧) between the control and label conditions may interfere with the perfusion signal.

The locations of the first frequency peak of X_*con*_(𝑧) and the zero-crossings of 𝐻(𝑧) can be calculated as Z_con_ in Eq. 1 and Z_0_ in Eq. 2, respectively. A solution for removing interferences was proposed that G_ratio_ can be adjusted according to Eq. 3 to overlap the first harmonic frequency peak with a neighboring k^th^ zero-crossing. This solution enables a slight change of gradient parameters within the optimal parameter range, and also automatically addresses the small interferences caused by X_*la*_(𝑧) as Z_la_ = 2 Z_con_. Because the first harmonic frequency peak resided within the first sidelobe of Labeling 0, two new labeling parameter sets, herein termed Labeling 1 and 2, were proposed by setting k = 2 and 3, respectively. Setting k = 1 was prohibited due to the relatively large sidelobes around the first zero-crossing, leading to unsatisfactory suppression of interference.

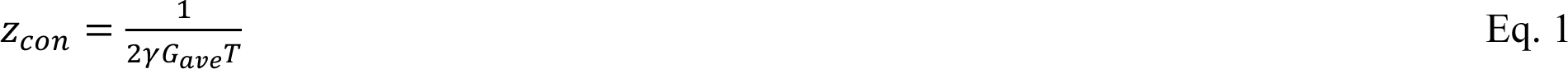

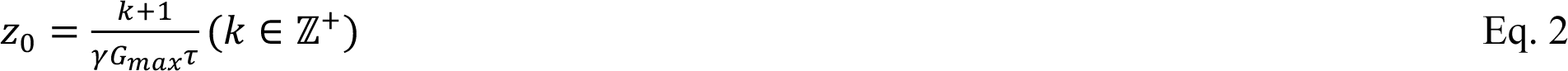

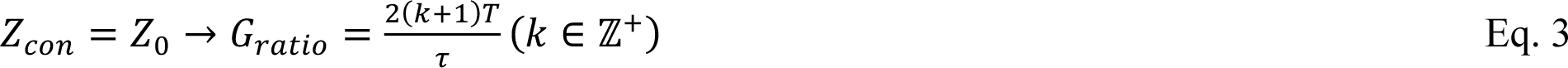

The above derivation is based on Fourier analysis, which ignores relaxations and is only legit in the low-tip-angle regime. To validate our analysis, we simulated Mz_con_(z), which denotes the longitudinal magnetization on static tissue (T1=1500 ms and T2=50 ms) immediately after pCASL labeling in control condition. The simulation was based on Bloch equation that modeled the RF pulses and gradients in pCASL labeling as a train of hard pulses with a step size of 10 µs (Dai et al., 2008).

The same simulation was repeated to optimize LE of pCASL labeling on arterial blood. Blood T1 and T2 values were assumed as 2087 ms and 68 ms, respectively, according to (Krishnamurthy et al., 2014; Zhang et al., 2013). LE was calculated as the average weighted by the flow contributions of different velocities in a cardiac cycle with the assumptions of laminar flow and the same pulsatile flow model for all feeding arteries (Zhao et al., 2017). As shown in Fig. 2 (a), following the work by (Wang et al., 2022), optimal gradient parameters were searched in a wide range of G_ave_ = [0.05:0.05:1.2] mT/m and G_ratio_ = [5:1:30] while fixing the other parameters, including T = 550 µs, 𝜏 = 300 µs, and the nominal flip angle (FA) = 15°. Then, a representative reduced B1 scaling of 0.5 and hardware limits of gradient strength = 80 mT/m and slew rate = 200 mT/m/ms were considered. Gradient parameters that violated hardware limits were excluded from the optimization. Potential optimal parameter sets were further analyzed in Fig. 2 (c-e) with a wide range of B0 (-200:20:200 Hz) and B1 scaling (0.2:0.05:1.2) values and compared to Labeling 0. The B1 and B0 ranges were determined based on a cohort including the values in 44 inflowing arteries from 11 subjects that was described in a previous study (Wang et al., 2022).

**Figure 2.**
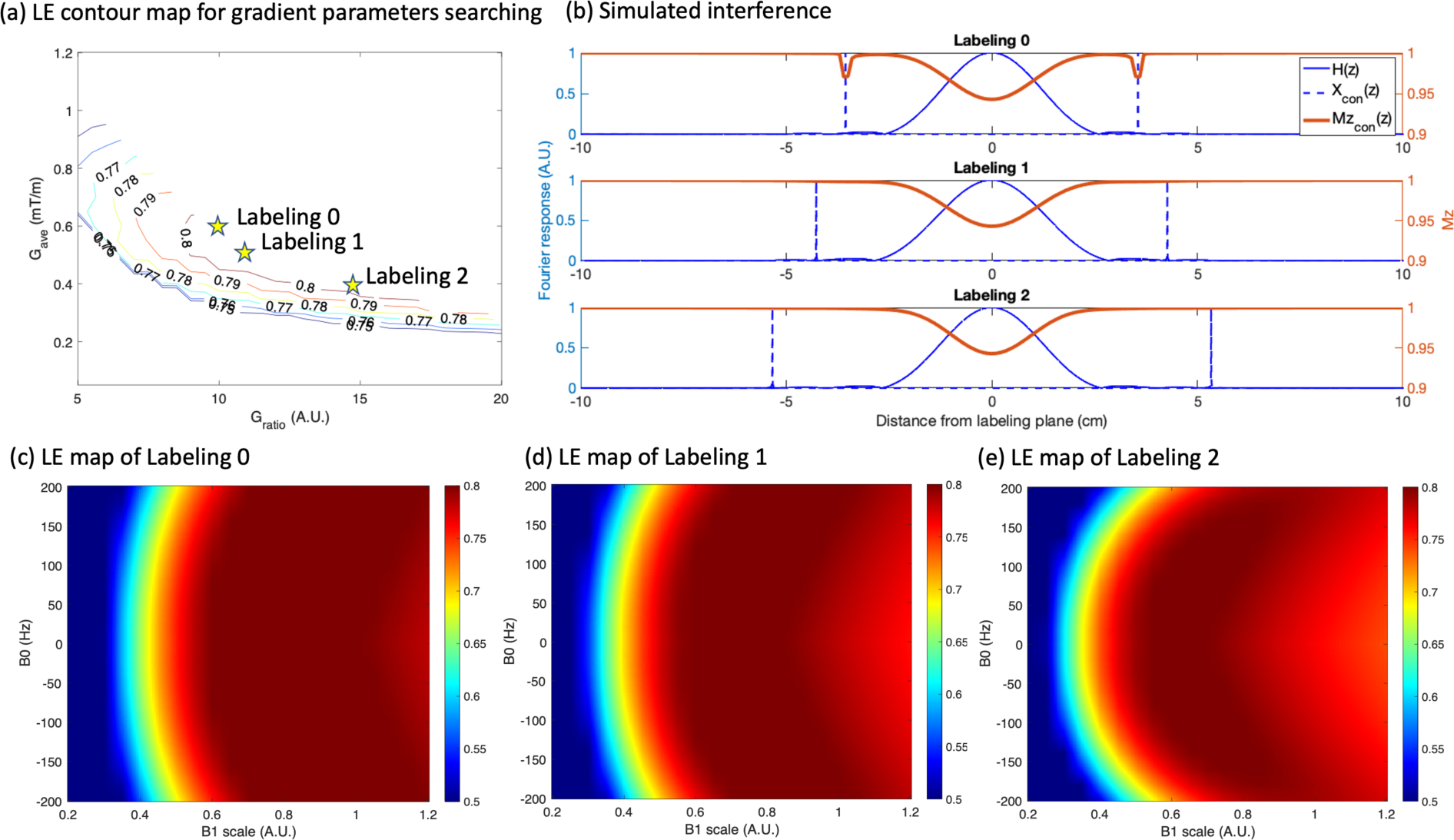
(a) LE contour map for gradient parameter searching. The original gradient parameters, Labeling 0, and two new gradient parameters, Labeling 1 and 2, satisfying *Eq. 3* were further analyzed. (b) Simulated interference of three pCASL labelings in the control condition. Compared to Labeling 0, the two new gradient parameters eliminate the interference by overlapping the first harmonic frequency peak with the zero-crossings. (c-e) Simulated LE maps under a range of B1 and B0 inhomogeneities. Despite of a similar average LE, new labeling parameters are more robust to low B1 but are slightly more sensitive to B0 offset.

### 2.2. Optimization of background suppression pulses

Four non-selective inversion pulses, including two optimal control-based pulses, MATPULSE and OPTIM pulse, and two traditional adiabatic pulses, Hyperbolic Secant (HS) and wideband uniform rate smooth truncation (WURST), were optimized and evaluated for the highest mean IE averaged across B0 offsets (±250 Hz) and B1 scalings (0.35-1.15). The optimization ranges were determined based on the B1 and B0 values in the pCASL imaging volume measured from a cohort of 9 subjects (see supplement section 2). Note IE was defined as -Mz(t)/Mz(0), where Mz(t) and Mz(0) represents the longitudinal magnetization before and after inversion (Garcia et al., 2005). Mz(t) was assumed to be negative and was set to 0 when a positive value occurs.

MATPULSE was initially designed for BS at 3T (3T pulse) using open-source software (Matson, 1994) with parameters of pulse duration = 10.24 ms, bandwidth = 1000 Hz, and peak amplitude = 10 µT. To adapt MATPULSE to the greater field inhomogeneities at 7T, the MATPULSE pulse was optimized based on parameter searching strategy by stretching pulse duration from 5 to 20 ms and peak amplitude from 5 to 20 µT. For HS and WURST, according to the definitions (see supplement section 3) (Kupce and Freeman, 1995; Silver et al., 1984), the parameter searching was expanded to include more pulse-specific parameters, such as μ (200-1000) and β (2-7) for HS and n (1-20) and k (250-1000) for WURST.

The OPTIM pulse was designed by means of ensemble-based time optimal control (Graf et al., 2022; Rund et al., 2018). Such a design alleviates the IE loss caused by the T2 relaxation during the pulse, and, thus, may achieve high performance at 7T. Detailed information can be found in supplement section 4. For the initialization of the optimization, random values of the RF pulse were assumed with a pulse duration of 10 ms. The box constraint of the RF amplitude was set to 20 µT. B1 and B0 ranges were slightly extended to 30-115% and ±280 Hz, respectively, in pulse design to achieve desirable performance in the target ranges, which were consistent with those of other pulses. The lower limit for B1 robustness was rather difficult to realize. Therefore, for a range of scales of 40% to 115%, the variable 𝜖 (see supplement section 4), which determines inversion quality, was set to 0.05, but increased to 0.1 for a range of 30% to 40%.

SAR levels were calculated as the time integral of the squared RF amplitudes, which was determined by pulse type, duration, maximum amplitude, and pulse-specific parameters. Relative SAR was calculated as the ratio of SAR of each pulse to that of 3T pulse. Pulses with different shapes were sorted by their relative SAR levels and shown in Fig. 3(a). For each pulse type, the parameter sets produced a lower mean IE at higher SAR levels were excluded.

**Figure 3.**
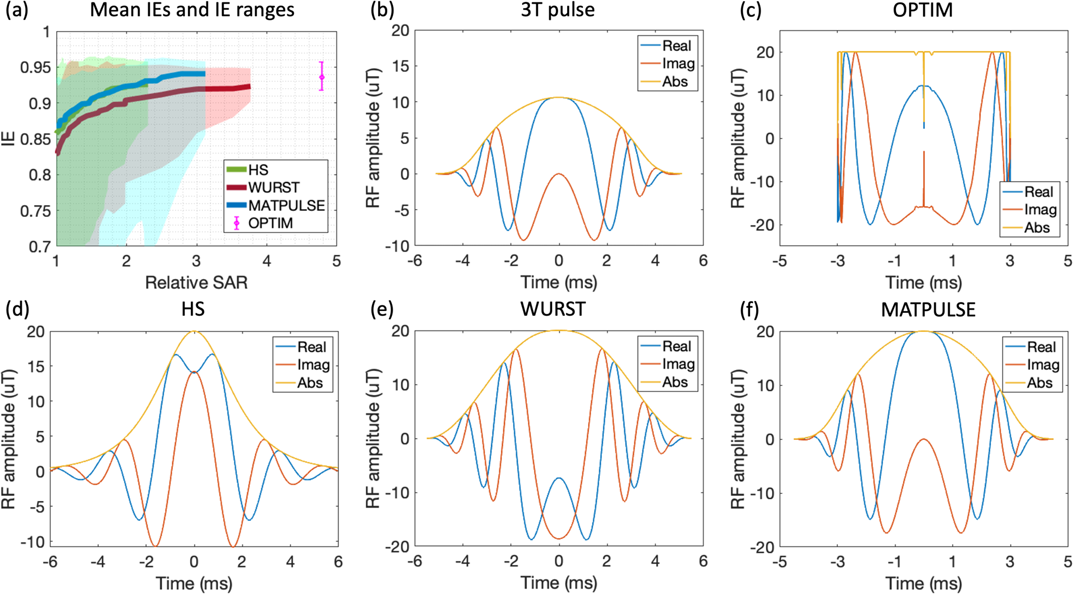
(a) Simulated IE of OPTIM and 3 BS pulses with different pulse-specific parameters and different relative SAR levels. The baseline of SAR levels was set by 3T pulse. The shaded area represents the IE range within the ranges of B1 scaling and B0 offset empirically defined at 7T. (b-f) 3T pulse and optimized inversion pulses for 7T. It was difficult to achieve a satisfying performance at a SAR level comparable to the 3T pulse. Among four pulses, OPTIM achieved the highest mean IE and min IE with a cost of 4.8-fold SAR. HS, WURST, and MATPULSE achieved their highest mean IE at relative SAR levels of 2.3, 3.7, and 3.1, respectively.

### 2.3. Optimization of 3D readout

As modeled by (Zuo et al., 2013), TFL-based perfusion signal Δ𝑆 at the j^th^ excitation can be described by Eq. 4,

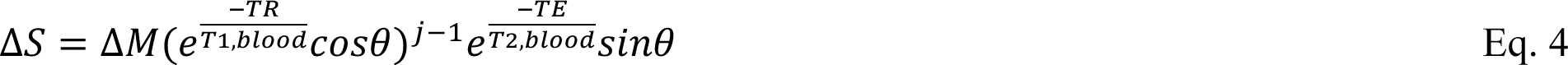

where 𝜃 is a small FA (<20^°^), and Δ𝑀 is the difference in longitudinal magnetization between label and control conditions at the start of the readout, which is related to physiological parameters, such as CBF and arterial transit time (ATT), and sequence parameters, such as labeling duration (LD) and post labeling delay (PLD). The model assumes the signal decays only with the T1 of blood, and indicates that the TFL-based perfusion signal decreases as more excitations/encoding-steps are applied and that the signal decays faster with a greater FA. Centric ordering was employed for phase encoding (PE) and partition directions to maximize SNR and &’/ flow contrast. Given j = 1, the signal acquired at the k-space center is 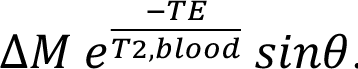. Consequently, the SNR of a ΔM map increases linearly with larger FA. On the other hand, the reduced signals in peripheral k-space may introduce image blurring. PE was looped inside the partition loop to avoid in-plane smoothness, and acceleration methods, including segmented readout and undersampling, were implemented to minimize through-plane blurring. 2D-CAIPIRINHA (Breuer et al., 2006) was implemented in TFL-pCASL sequence to minimize aliasing artifacts. Fig. 4 (a) illustrates the k-space sampling pattern and the change of perfusion signals during data acquisition.

**Figure 4.**
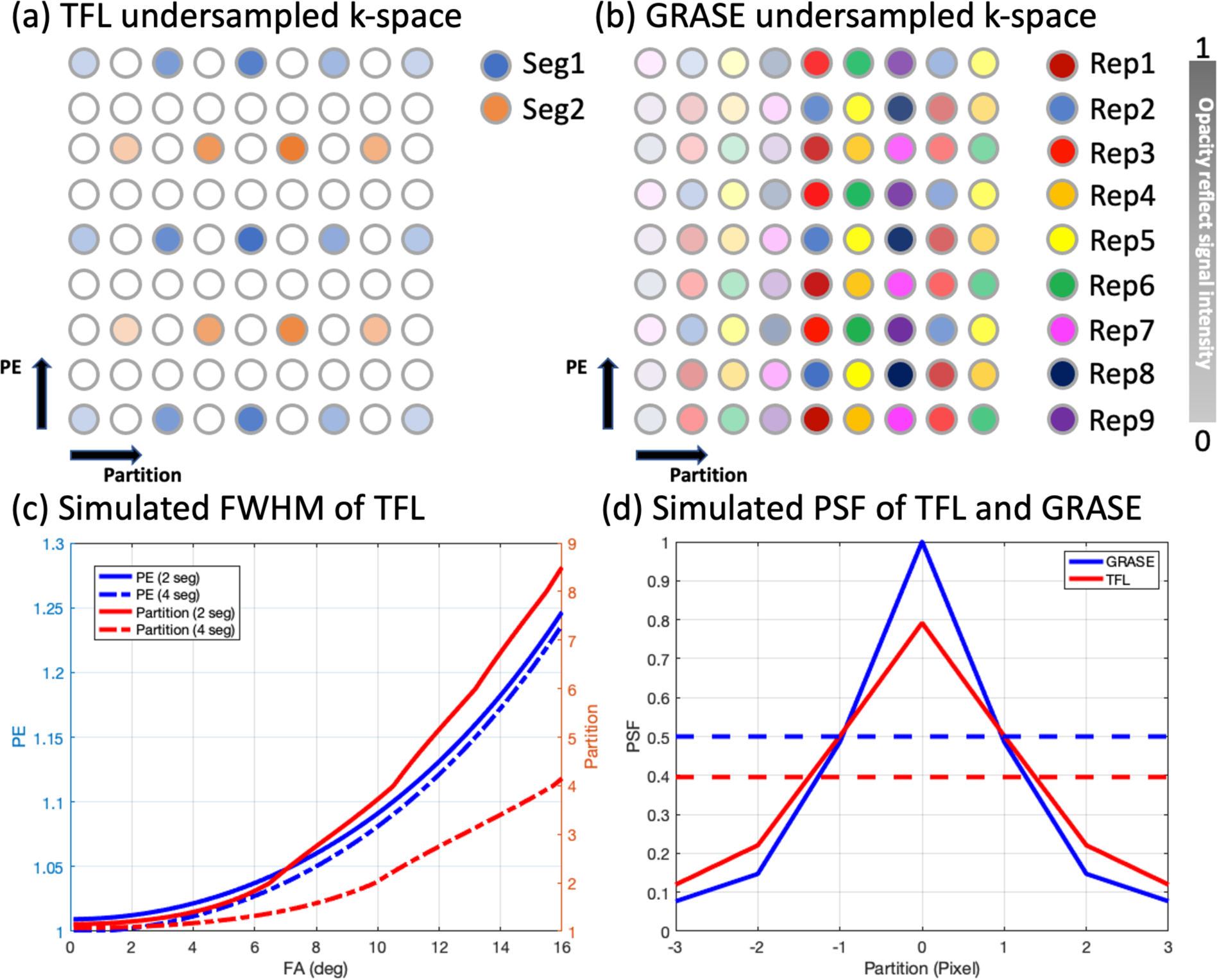
(a, b) k-space undersampling pattern of TFL-pCASL and GRASE-pCASL. The opacity change indicates the decay of the perfusion signal. TFL-pCASL was accelerated by 2 segments and 2D-CAIPIRINHA undersampling with R=2×2. The single shot GRASE-pCASL was undersampled by time-dependent 2D-CAIPIRINHA with R=3×3. 9 repetitions of control/label pairs have interleaved k-space samples. (c) FWHM estimated from the simulation of PSF with varying FAs and N_seg_s. Blurring was minimal in the PE direction but increased with increased FA and decreased the number of segments in the partition direction. (d) Simulated PSFs of GRASE and TFL. The amplitudes of PSFs reflect SNR of two methods. PSF of GRASE-pCASL showed a 20% higher SNR and 0.7 pixels narrower width than that of TFL-pCASL.

As a comparison, GRASE-based perfusion signal Δ𝑆_)6789_ at k^th^ spin echo and l^th^ gradient echo can be described by Eq. 5,

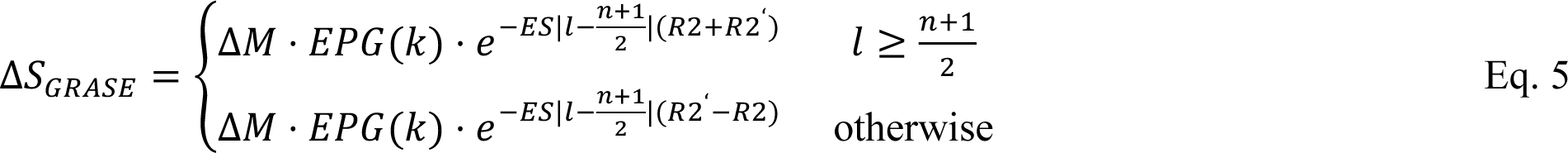

where n is EPI factor (odd number only), ES is the echo spacing in EPI, R2=1/T2, R2’=1/T2*-1/T2, and EPG(k) is the simulated k^th^ spin echo signal using extended phase graphs (EPG) (Carr-Purcell-Meiboom-Gill (CPMG) and FA of 120°) (Hargreaves, 2022; Weigel, 2015). The perfusion signal decays quickly due to short T2 and T2* at 7T, which requires a fast acquisition.

As shown in Fig. 4 (b), a single shot GRASE sequence was accelerated by a time-dependent 2D-CAIPIRINHA R-fold undersampling (Spann et al., 2020). The sequence consisted of an integer multiple of R label/control pairs, in which the k-space samples were interleaved within every R pairs. A joint reconstruction of R label/control pairs yielded R individual ΔM maps with reduced aliasing artifacts, as TGV-regularization improves SNR by exploiting temporal-spatial smoothness (Spann et al., 2017).

Point spread function (PSF) and associated full width half max (FWHM) of a ΔM map were simulated for TFL-pCASL as a function of FA and the number of segments (N_seg_) based on the signal evolution described in Eq. 4, the sampling fashions stated above, and the same imaging parameters as in-vivo scans presented in section 2.4. For comparison, the simulation was repeated for GRASE-pCASL using the in-vivo imaging parameters shown in section 2.4. In addition to T1 and T2 of arterial blood mentioned in section 2.1, T2* of 30 ms (Peters et al., 2007) was also assumed. The amplitudes of PSFs were normalized by SNR, taking into account the signal intensity acquired at the k-space center, bandwidth, and undersampling rate (R). Effect of regularization in image reconstruction was not considered.

### 2.4. In-vivo Experiments

TFL-pCASL was performed with a bandwidth of 490 Hz/pixel to maximize SNR while maintaining a short TE of 1.5 ms and echo spacing (ES) of 4 ms. For each subject, T1w images were acquired by using product MP2RAGE sequence, and time-of-flight (TOF) angiography images were acquired to place the labeling plane at the C1 segment (Bouthillier classification) of the internal carotid arteries (see supplement section 5). pCASL labeling parameters were: balanced scheme, T = 550 µs, 𝜏 = 300 µs, G_ave_ = 0.4 mT/m, and G_ratio_ = 14.67, nominal FA = 15°, LD = 1 s, PLD = 2 s. Two OPTIM BS pulses were applied at 427 and 1530 ms after labeling to suppress static tissue signals (T1 = 1-2 s) to 10% at the beginning of the readout. Other imaging parameters were: resolution = 2×2×4 mm^3^, FOV = 224×192×112 mm^3^, matrix size = 112×96×28 including 4 oversampled slices, 2D-CAIPIRINHA with R=2×2, N_seg_ = 2, FA = 8^°^, readout duration = 1344 ms, effective TR (TR_eff_) = 7 s (2.6 s dead time (TD)), 50 measurements, including an M0 image, acquired in 11 mins 40 s. A fully sampled calibration scan was acquired separately without undersampling in 20 s for estimation of GRAPPA weights. Unless otherwise stated, the parameters in this protocol were used as default.

Single slice 2D TFL sequences with and without a BS pulse were used to obtain whole brain IE maps of the BS pulses in axial, sagittal, and coronal views (see supplement section 6). The imaging parameters were: resolution = 2.2×3 mm^2^, FOV=210×192mm^2^, slice thickness=5 mm, matrix size=96×48, linear ordering, 6/8 partial Fourier, slice thickness=5 mm, FA=8°, TE=1.38 ms, ES=3.3 ms, readout duration=150 ms, and TR_eff_=10 s that allows a full relaxation.

GRASE-pCASL was performed using the same pCASL labeling and BS scheme as TFL-pCASL. The imaging parameters were: resolution = 2.1×2.1×4 mm^3^, FOV = 200×200×120 mm^3^, matrix size = 112×93×30 including 6 oversampled slices, bandwidth = 2170 Hz/pixel, ES = 0.53 ms, TE = 22 ms, 2D-CAIPIRINHA with R=3×3, EPI factor = 31, turbo factor = 10, single shot, FA=120^°^, readout duration = 220 ms, TR_eff_ = 8 s (TD = 4.7 s), 63 measurements, including an M0 image, were acquired in 8 min 40 s. The imaging parameters were chosen to best match those of TFL-pCASL as well as to balance TE and required number of spin echoes according to our previous study (Shao et al., 2020).

SMS-TFL-pCASL was compared to 3D TFL-pCASL with the default protocol. SMS-TFL-pCASL used the same labeling parameters as 3D TFL-pCASL, and the following imaging parameters: resolution = 2×2×4 mm^3^, FOV = 224×192×100 mm^3^, matrix size = 112×72×25, 6/8 partial Fourier along PE, linear ordering (k-space center was acquired at the 24^th^ excitation), bandwidth = 490 Hz/pixel, no slice gap, ES=3.3 ms, SMS slice group acquisition = 245 ms, SMS factor = 5, 1/4 FOV shift, FA = 8^°^, TR_eff_ = 4.3 s (TD = 0 s), no BS, 122 measurements, including an M0 image, acquired in 8 mins 45 s. TR_eff_ of 3D TFL-pCASL in this comparison was shortened to 5.2 s (TD = 0.9 s) to match the total scan time.

Comparison experiment of 7T and 3T was conducted using the default TFL-pCASL protocol with adjustments. At 3T, a commonly used set of balanced pCASL parameters include T = 920 µs, 𝜏 = 500 µs, G_ave_ = 0.6 mT/m, G_ratio_ = 10, LD = 1.8 s, PLD = 2 s, and a fixed labeling plane that is 90 mm away from the imaging center. Other changes at 3T included adjusted BS timing at 60 and 1500 ms following label, shortened TR_eff_ = 5.2 s (TD = 0 s), and total scan time = 8 min 40 s. At 7T, TR_eff_ was adjusted to the shortest possible value (on average 5.6 s) for each subject based on the maximal allowed SAR.

There were eight MRI experiments to evaluate: 1) 3 labeling parameter sets (3 subjects), 2) BS pulses (1 subject), 3) N_seg_ of 2 and 4 (same acquisition time, 1 subject), 4) FA of 4, 8, and 12° (1 subject), 5) comparison of TFL- and GRASE-pCASL (3 subjects), 6) test-retest repeatability of TFL- and GRASE-pCASL (1 day apart, 5 subjects), 7) comparison of 3D TFL- and 2D SMS-TFL-pCASL (1 subject), 8) comparison of 3T and 7T (5 subjects)

In total, nineteen healthy subjects (8 females, 28.1±5.0 y/o) underwent MRI experiments on the investigational pTx part of a 7T Terra system (Siemens Medical Systems, Erlangen, Germany) with an 8Tx/32Rx head coil (Nova Medical, Cambridge, MA, USA) on the first level of SAR, and a 3T Prisma system (Siemens Healthineers, Erlangen, Germany) using a 32-channel head coil under the approval of a local IRB. All subjects provided written informed consents. At 7T, the RF-shimming mode was TrueForm (Nistler et al., 2007), which uses 45° phase increment for each adjacent transmit channel to mimic the single transmit circularly polarized (CP) coil. B0-shimming was performed over the imaging volume with an extended lower boundary to include labeling plane (see supplement section 5).

### 2.5. Data analysis

All numerical simulations and optimizations were performed in MATLAB (The Mathworks Inc., Natick, USA). An in-house GUI-based toolbox was also developed in MATLAB for offline image reconstruction of TFL-pCASL. GRAPPA (Griswold et al., 2002) with a kernel size of 7×4×2 was adapted for 2D-CAIPIRINHA undersampling pattern for image reconstruction. For GRASE-pCASL, an open-source toolbox (“https://github.com/IMTtugraz/AVIONIC,” 2022; Schloegl et al., 2017) was used for TGV-regularized reconstruction with default parameters (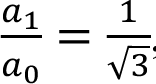, w=0.9, and ß=7) and λ = 10 as suggested by (Spann et al., 2020). M0 and label/control images of GRASE-pCASL were reconstructed separately. SMS-TFL-pCASL images were reconstructed offline using an in-house MATLAB toolbox of Slice-GRAPPA with 3×3 kernel (Setsompop et al., 2012).

Cerebral blood flow (CBF) was quantified using the single-compartment kinetic model adapted for TFL-pCASL by (Zuo et al., 2013) as described in Eq. 6,

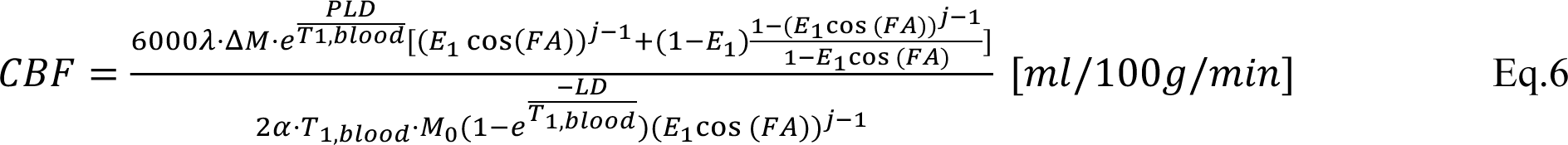

where 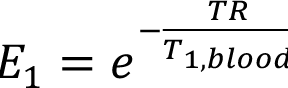, j was set to 1 for centric ordering, 𝜆 is the brain/blood partition coefficient, which was assumed as 0.9 ml/g, 𝛼 (=LE*IE^2^) is the labeling efficiency. The definitions and values of other parameters were as defined above. For GRASE-pCASL, the standard pCASL single-compartment kinetic model was used (Alsop et al., 2015). At 3T, LE and IE of BS were assumed as 0.85 and 0.95, respectively, resulting in a 𝛼 = 0.76 according to (Alsop et al., 2015). At 7T, 70% 𝛼 at 3T was assumed for 𝛼 at 7T based on our 3T and 7T comparison results presented in section 3.6.

ΔM maps were obtained by subtracting label images from control images. SNR was evaluated as the ratio of the mean to the spatial noise standard deviation (SD) of the ΔM maps. According to (Feinberg et al., 2013), the spatial noise SD was calculated from a difference image obtained by subtracting the average of all odd-indexed ΔM maps from the average of all even-indexed ΔM maps. tSNR was evaluated as the mean ratio of the mean to the temporal noise SD across all repetitions of ΔM maps on each voxel. Through-plane blurring of a ΔM map was quantified using blur quality metric (BQM) (Crete et al., 2007), which is a value between 0 and 1, and a larger value indicates an image is more blurred. Test-retest repeatability was evaluated using within-subject coefficient of variance (wsCV) (Bland and Altman, 1996) and intraclass correlation coefficient (ICC) (McGraw and Wong, 1996) based on average CBF values of five subjects.

GM, WM, and brain masks were segmented on T1w images using SPM12 (The Wellcome Centre for Human Neuroimaging, UCL Queen Square Institute of Neurology, London, UK). For all comparison studies, T1w images and associated masks were individually coregistered to ASL images acquired with different methods. ΔM, SNR, tSNR, and BQM were measured in brain masks, while ΔM/M0, CBF, wsCV, and ICC in individual masks were reported. Measurements including multiple subjects were reported in the form of mean ± SD across subjects. In comparison of TFL- and GRASE-pCASL, SNR measurements of GRASE-pCASL were corrected to match the acquisition time of TFL-pCASL.

## 3. Result

### 3.1. Optimization of pCASL labeling

Figure 2 (a) shows the simulation result of LE contour map. The LE contour was open-ended in the top right corner because of exceeded slew rate. According to the contour map, two candidates, Labeling 1 (G_ave_ = 0.5 mT/m and G_ratio_ = 11) and Labeling 2 (G_ave_ = 0.4 mT/m and G_ratio_ = 14.67), that conformed to the requirement defined in Eq. 3 were selected from a set of optimal parameters achieving LE greater than 0.8.

Figure 2 (b) show 𝐻(𝑧) (solid blue), X_*con*_(𝑧) (dotted blue), and 𝑀𝑧_*con*_(𝑧) (red). Mz_con_(z) was expected to follow the profile of 1-𝐹_*con*_(𝑧) because 𝐹_*con*_(𝑧) represent excitation profile in transverse plane. Interferences in Labeling 0 were eliminated by exactly placing Z_con_ at the second and third Z_0_. Although the interferences only led to a magnetization difference of 0.03 on static tissue, the perfusion signal, typically less than 1% of the tissue signal, can still be overwhelmed. Between Labeling 1 and 2, the latter showed better suppression of interferences.

Figure 2 (c-e) characterize the LE of two candidate parameter sets in the presence of B1/B0 field inhomogeneities. From Labeling 0, 1 to 2, the decreasing G_ave_ improved the robustness of LE to low B1, while the increasing G_ratio_ increased the sensitivity of LE to B0 inhomogeneities. Within the optimization range, the same simulated LE of 0.73 was found for Labeling 0, 1, and 2.

Figure 5 presents in-vivo ΔM/M0 maps from 3 subjects. With Labeling 0, strong interferences of dark bands (red arrows) can be observed in the bottom slices. These dark bands disappeared in the maps of Labeling 1 and 2, suggesting that the proposed labeling parameter sets minimized interferences. Compared to Labeling 1, Labeling 2 achieved a slightly better suppression because of a reduced amplitude of higher order sidelobes. In addition, the perfusion signal was high and uniform in ΔM/M0 maps without noticeable asymmetrical labeling effects attributed to the robustness of all labeling parameter sets to field inhomogeneities. ΔM/M0 in gray matter (GM) measured from 3 subjects were 0.24 ± 0.04, 0.24 ± 0.03, and 0.26 ± 0.06% for Labeling 0, 1, and 2, respectively. White matter (WM) ΔM/M0 values were 0.15 ± 0.02, 0.16 ± 0.01, and 0.16 ± 0.03% for Labeling 0, 1, and 2, respectively.

**Figure 5.**
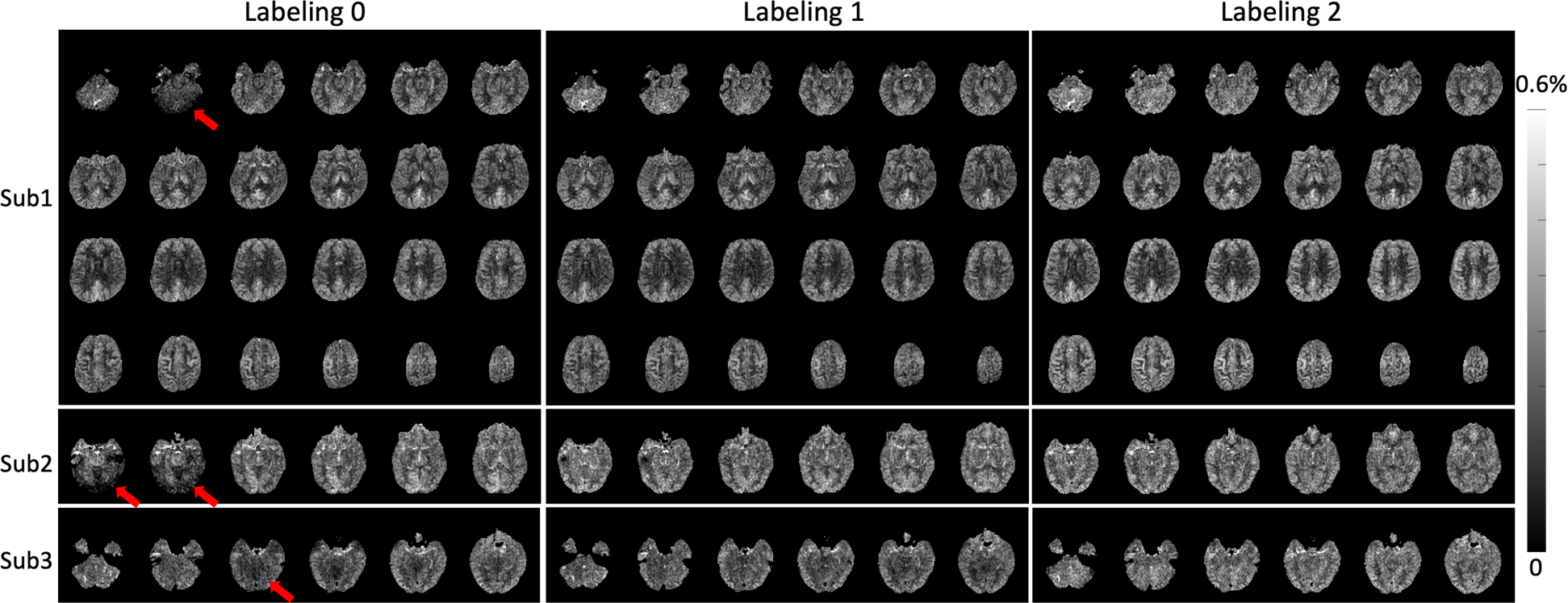
In-vivo axial slices of ΔM/M0 maps acquired from 3 subjects. Only the bottom 6 slices were shown for subject 2 and 3. Severe interference (red arrows) observed in Labeling 0 were eliminated by Labeling 1 and Labeling 2. Compared to Labeling 1, Labeling 2 showed better performance, which was consistent with the theoretical analysis and simulation results from *Fig. 2* (b). From in-vivo images acquired on the three subjects, no significant difference in LE among the three labeling parameter sets was found, as evidenced by comparable ΔM/M0 values.

### 3.2. Optimization of background suppression pulses

Figure 3 (a) presents mean IEs and the associated ranges as the shaded area achieved by the four BS pulses as a function of relative SAR from the simulation. 3T pulse yielded compromised performance at 7T with a mean IE of 0.87 and minimal IE of 0.4. With the double BS scheme, this may reduce the perfusion signal by 25%-80%. Generally, it was difficult to achieve acceptable performance with a relative SAR < 2. At 2.3-fold SAR, HS achieved a mean IE of 0.92 and a minimal IE of 0.87. At 3.1-fold SAR, MATPULSE achieved a mean IE of 0.94 and a minimal IE of 0.86. At 3.7-fold SAR, WURST achieved a mean IE of 0.92 and a minimal IE of 0.90. Among all tested pulses, OPTIM achieved the highest mean IE of 0.94 and the highest minimal IE of 0.92 at the cost of 4.8-fold SAR. In practice, the choice of different BS pulses can be determined by the maximal allowed SAR and acceptable scan time. The optimized pulses are presented in Fig. 3 (b-f), and the parameters are listed in supplement section 3.

Figure 6 compares the optimized pulses with 3T pulse in IE maps and 𝛥M/M0 maps in vivo. It can be seen that significantly higher and more uniform IEs were achieved by the optimized pulses compared to 3T pulse. For 3T pulse, MATPULSE, HS, WURST, and OPTIM, mean IE were 0.85, 0.89, 0.89, 0.91, and 0.92, respectively, and coefficients of variance (CV) were 0.12, 0.06, 0.08, 0.08, and 0.04, respectively. Among optimized pulses, OPTIM offered the best performance (9.4% IE increase and 66.6% CV decrease compared to 3T pulse). In addition, a critical deficiency of IE in bottom slices and temporal lobes (red arrow) was caused by the sensitivity of 3T pulse to field inhomogeneities in inferior regions. The low IE may raise the concern of substantial loss of perfusion signal and shortening of efficient LD. From 𝛥M/M0 maps, the perfusion signal was significantly higher for OPTIM compared to 3T pulse. As listed in Table 1, compared to 3T pulse, OPTIM increased GM 𝛥M/M0 by 33.3% from 0.21 to 0.28% and tSNR by 23.5% from 0.85 to 1.05.

**Figure 6.**
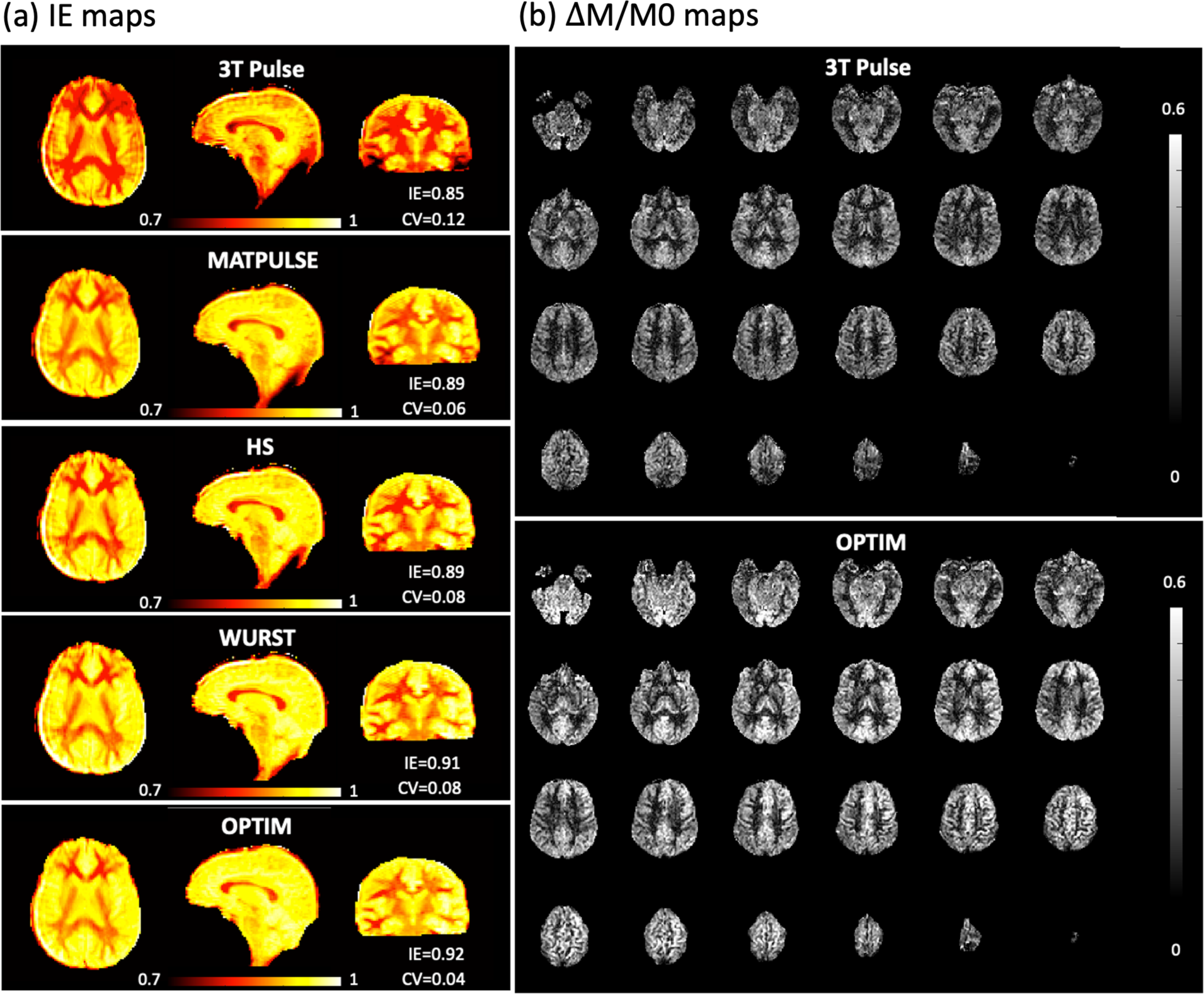
(a) In-vivo IE maps acquired with a single slice (in all 3 views) 2D TFL readout immediately following a BS pulse. Optimized pulses achieved higher IE and lower CV than 3T pulse, where OPTIM offers the best performance. (b) In-vivo 𝛥M/M0 maps acquired with OPTIM and the 3T pulse. OPTIM offers 33.3% higher perfusion signal and 23.5% higher tSNR in GM than 3T pulse.

**Table 1.** Measurements of perfusion signal and performance metrics with different labeling parameters, BS pulses, TFL and GRASE readout

| | $\Delta M/M0$ (%) in GM | $\Delta M/M0$ (%) in WM |
| --- | --- | --- |
| Labeling 0 | $0.24 \pm 0.04$ | $0.15 \pm 0.02$ |
| Labeling 1 | $0.24 \pm 0.03$ | $0.16 \pm 0.01$ |
| Labeling 2 | $0.26 \pm 0.06$ | $0.16 \pm 0.03$ |
| | $\Delta M/M0$ (%) in GM | tSNR |
| 3T pulse | 0.21 | 0.85 |
| OPTIM | 0.28 | 1.05 |
| | BQM* | $\Delta M$ (a.u.) |
| FA = 4° | 0.24 | 0.37 |
| FA = 8° | 0.30 | 0.69 |
| FA = 12° | 0.34 | 1 |
|  | BQM* | SNR |
| N<sub>seg</sub> = 2 | 0.30 | 2.63 |
| N<sub>seg</sub> = 4 | 0.27 | 1.26 |
|  | BQM* | SNR |
| TFL-pCASL | $0.32 \pm 0.00$ | $3.75 \pm 0.08$ |
| GRASE-pCASL | $0.30 \pm 0.04$ | $5.37 \pm 1.38$ |
|  | CBF GM (ml/100g/min) | CBF WM (ml/100g/min) |

|  | Test | Retest | Test | Retest |
| --- | --- | --- | --- | --- |
| TFL-pCASL | 35.8±2.7 | 35.1±2.2 | 20.5±1.4 | 20.5±1.9 |
| GRASE-pCASL | 38.9±2.2 | 36.3±3.6 | 24.1±2.4 | 21.7±2.6 |

|  | SNR | tSNR |
| --- | --- | --- |
| 3D TFL-pCASL | 2.45 | 1.07 |
| SMS-TFL-pCASL | 1.33 | 0.31 |

| | SNR | $\Delta M/M0$ (%) in brain/GM/WM |
| --- | --- | --- |
| 7T | 2.24±0.28 | 0.20±0.02 / 0.24±0.02 / 0.11±0.02 |
| 3T | 1.58±0.27 | 0.32±0.05 / 0.38±0.06 / 0.19±0.04 |

\* The blur quality metric (BQM) is a value between 0-1, and a larger value indicates an image is more blurred.

### 3.3. 3D readout

Figure 4 (c) demonstrates the change of FWHM as a function of FA in TFL-pCASL, where FWHM increased with larger FA and reduced N_seg_. Minimal blurring was introduced in the PE direction with an FWHM less than 1.1 regardless of N_seg_ tested. Nevertheless, more blurring in partition direction was demonstrated. At FA of 4°, 8°, and 12°, FWHMs were 1.3, 2.7, and 5.1 pixels, respectively, for N_seg_ of 2, which were reduced to 1.2, 1.6, and 2.7 pixels, respectively, by increasing N_seg_ to 4. Since SNR increases linearly with larger FA, there is a trade-off between SNR and through-plane blurring. Although a greater N_seg_ eased this trade-off, artifacts related to motion or physiological fluctuation between segments may arise. Therefore, according to this simulation, a TFL readout with N_seg_ of 2 and FA of 8° may be recommended.

Figure 4 (d) shows the PSFs simulated for ΔM maps acquired using TFL and GRASE readouts in the partition direction. No significant blurring was found in PE direction for both methods (see supplement 7). Comparing the two PSFs, the amplitude of TFL was 20% lower than that of GRASE, and the FWHM of TFL was 2.7 pixels, which was 0.7 pixel wider than that of GRASE. Simulation results suggested that, with a 9-fold acceleration and the advanced TGV reconstruction, GRASE may achieve higher SNR and less through-plane smoothness than TFL.

Figure 7 (a) and (b) show in-vivo TFL-pCASL ΔM maps in axial, coronal, and sagittal views acquired with different FAs and N_seg_s. Gray scales were adjusted to visualize ΔM maps with different signal levels. The image was noisy, and GM/WM contrast was plain for FA of 4°. By increasing FA to 8° and 12°, signal amplitudes were doubled and tripled, respectively, resulting in improvements in SNR and GM/WM contrast. As listed in Table1, the ΔM values of 4° and 8° relative to that of 12° were 0.37 and 0.69, respectively. However, through-plane blurring was aggravated by increased FA, which was demonstrated by the coronal and sagittal views of the ΔM maps in Fig. 7 (a). BQMs measured for FA of 4°, 8°, and 12° were 0.24, 0.30, and 0.34, respectively. Similarly, from Fig. 7 (b), more through-plane blurring can be seen in the ΔM maps with N_seg_ = 2 than that with N_seg_ = 4, and the BQMs were 0.32 and 0.28 for N_seg_ of 2 and 4, respectively. However, increasing N_seg_ introduced ringing artifacts (red arrows) that might be related to the motion or physiological fluctuation. In addition, the ΔM map with N_seg_ of 4 was noisier than the ΔM map with N_seg_ of 2 because the number of repetitions was halved. As listed in Table 1, SNR were 2.63 and 1.26 for N_seg_ of 2 and 4, respectively.

**Figure 7.**
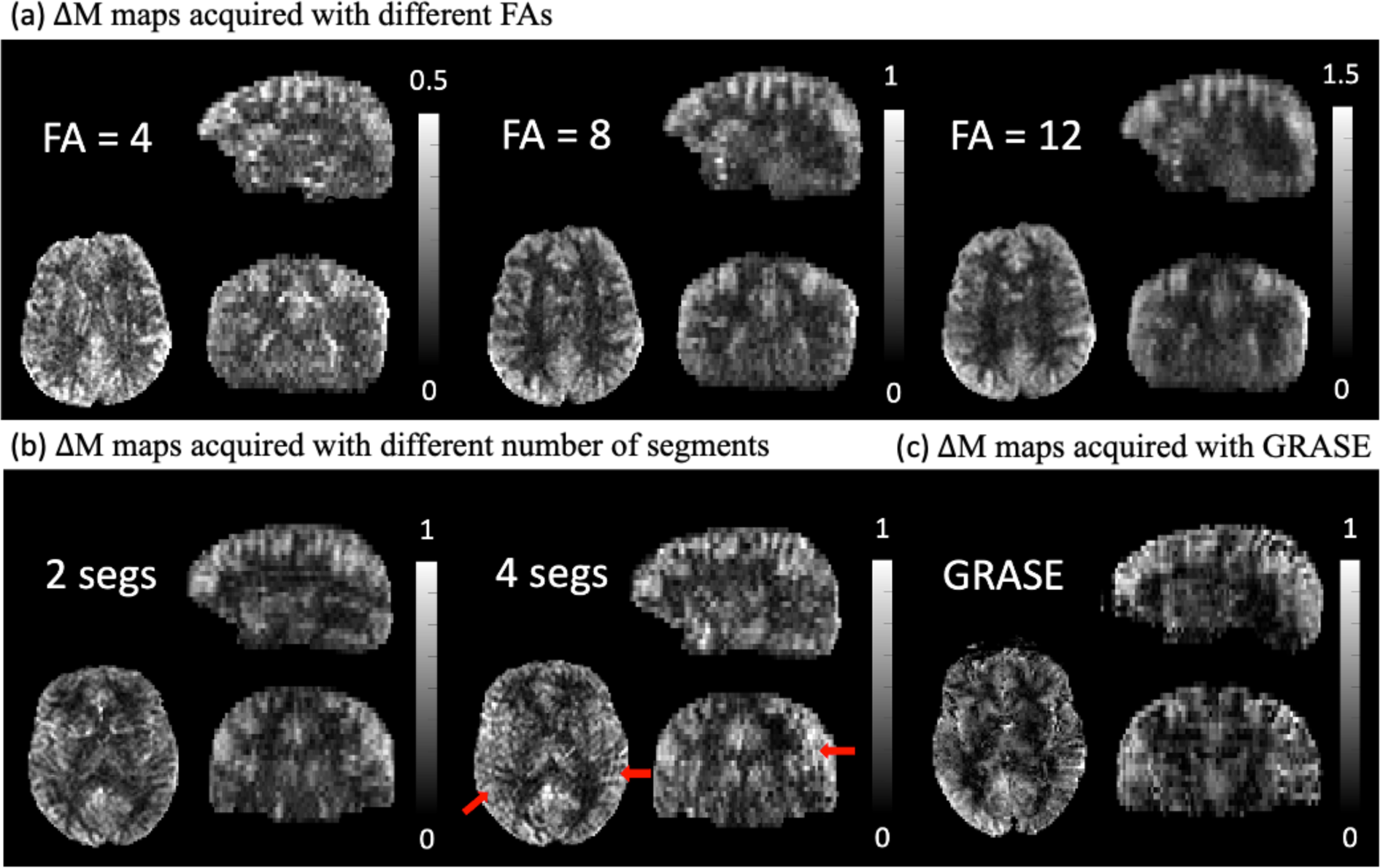
(a) ΔM maps acquired with different FAs demonstrating a trade-off between perfusion signal intensity and spatial blurring in the partition direction (N_seg_ = 2). (b) ΔM maps acquired with different numbers of segments (FA = 8°). Despite increased sharpness in the partition direction, the ΔM map acquired with 4 segments had half number of measurements and was prone to motion-induced artifacts. (c) The ΔM map acquired with GRASE on the same subject as (b). Although SNR and sharpness of GRASE-pCASL were comparable to TFL-pCASL, GRASE-pCASL suffered from distortion and intensity variations that may compromise its reliability.

TFL- and GRASE-pCASL are compared in in-vivo ΔM maps in Fig. 7 (b,c) and CBF maps in Fig. 8. In Fig. 7 (b,c), GRASE-pCASL showed a higher SNR and less through-plane smoothness compared to TFL-pCASL with N_seg_ = 2, and a higher SNR but smoother coronal and sagittal views compared to TFL-pCASL with N_seg_ = 4. Fig. 8 reveals distortions (red arrows) in regions, such as lower slices and frontal lobes, in GRASE-pCASL, which were not observed in TFL-pCASL images. Supplement section 8 shows ΔM/M0 maps overlaid on T1w images, where TFL-pCASL matched T1w images better than GRASE-pCASL, especially in frontal and temporal regions. Besides, CBF maps in Fig. 8 show abnormal intensity variations in GRASE images (yellow arrows), probably due to susceptibility artifacts and unresolved aliasing artifacts.

**Figure 8.**
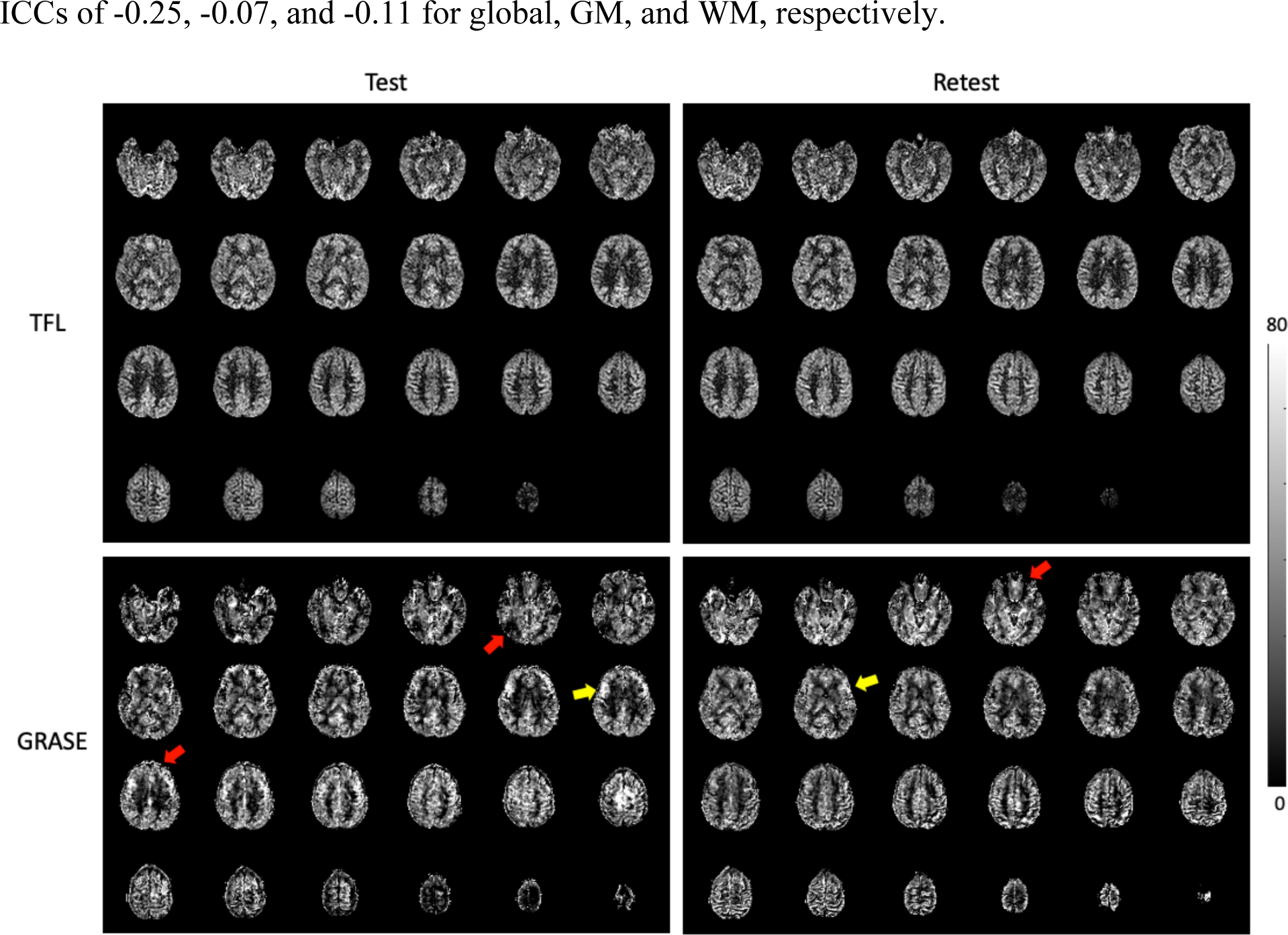
CBF (ml/100g/min) maps of a representative subject in two visits using TFL- and GRASE-pCASL.

Some bright vascular signals can be observed in TFL-pCASL ΔM/M0 maps due to arterial transit effects. Table 1 reports that the SNRs were 3.75±0.08 and 5.37±1.38 and the BQMs were 0.32±0.01 and 0.30±0.03 for TFL and GRASE, respectively. Generally, although GRASE-pCASL offered better SNR and BQM measurements, distortion-free pCASL images with whole-cerebrum coverage and better image quality were achieved by 3D TFL readout. Many anatomical features can be easily discerned, such as orbitofrontal cortex, choroid plexus, white matter, and cortical gyri. Higher resolution (2 mm isotropic) is possible with reduced SNR (see supplement section 9)

### 3.4. Test-retest repeatability

Figure 8 shows CBF maps of a representative subject in two visits using TFL- and GRASE-pCASL. Measured CBF values are listed in Table 1. CBF maps of other subjects and scatter plots of test-retest results of TFL- and GRASE-pCASL are listed in supplement section 10. TFL-pCASL provided a good to excellent repeatability. For brain, GM, and WM, wsCVs were 3.9%, 4.8%, and 3.6%, and ICCs were 0.64, 0.47, and 0.80, respectively. However, the test-retest repeatability of GRASE-pCASL was compromised with wsCV of 9.0%, 8.9%, and 12.8%, and ICCs of -0.25, -0.07, and -0.11 for global, GM, and WM, respectively.

### 3.5. Comparison of 3D and 2D SMS TFL readouts

Figure 9 shows ΔM/M0 maps acquired by using SMS-TFL-pCASL and 3D TFL-pCASL. CBF maps were shown in supplement section 11. Perfusion signal of SMS-TFL-pCASL decayed in the slice groups (columns) acquired at longer PLDs, while consistent perfusion signal and better image quality in all slices were observed with 3D TFL-pCASL. SNR was 1.33 and 2.45 for SMS-TFL-pCASL and 3D TFL-pCASL, respectively. In addition, substantial temporal noise (see supplement section 11 for SD maps) arose in SMS-TFL-pCASL due to the lack of BS. tSNRs were 0.31 and 1.07 for SMS-TFL-pCASL and 3D TFL-pCASL, respectively. In addition, as shown in inset of Fig. 9, slice cross-talk was observed in sagittal and coronal views of SMS-TFL-pCASL.

**Figure 9.**
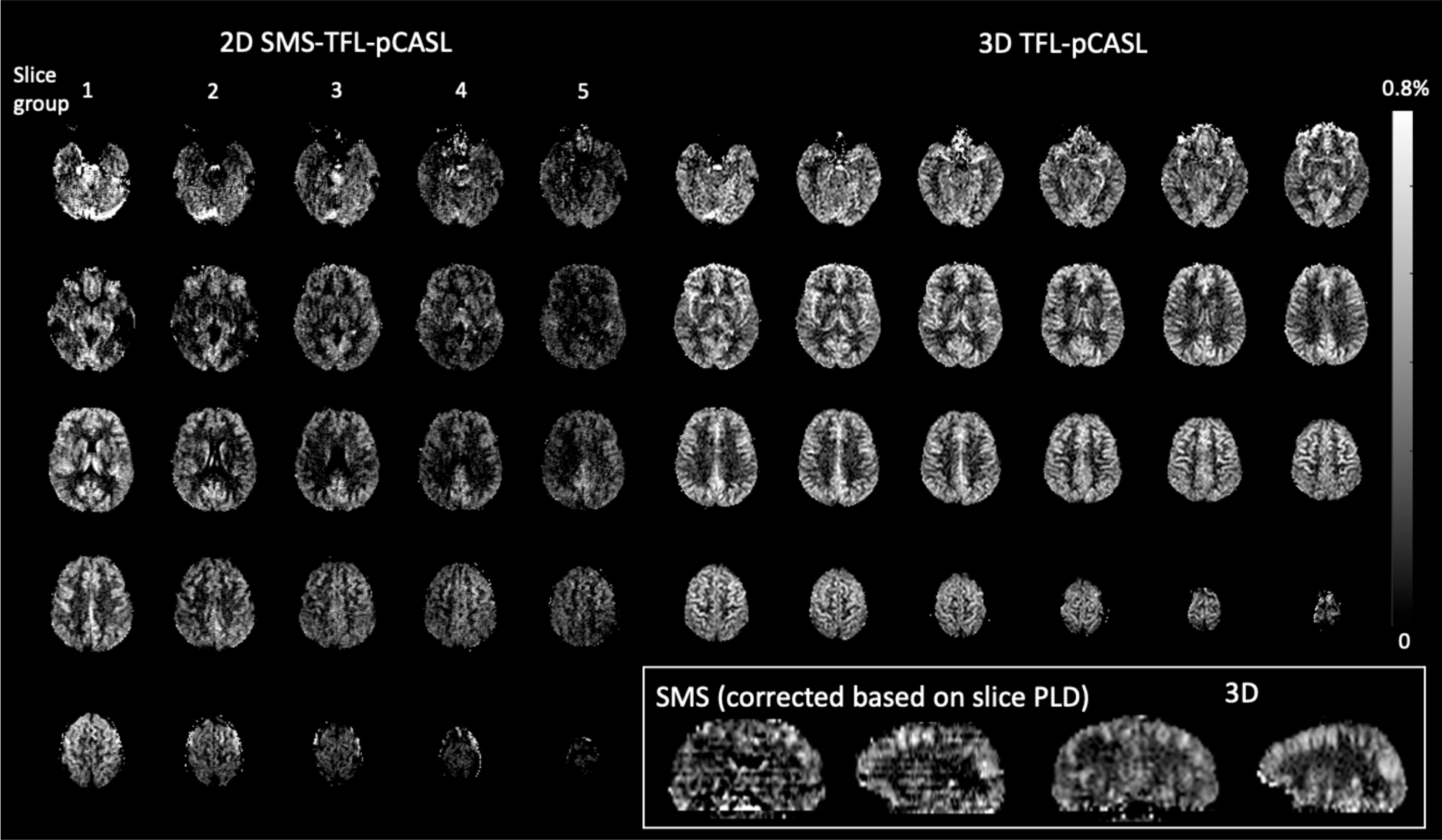
𝛥M/M0 maps for a comparison of 2D SMS-TFL-pCASL and 3D TFL-pCASL. The inset shows the coronal and sagittal views of the two methods.

### 3.6. Comparison of 3T and 7T TFL-pCASL

Figure 10 shows the CBF maps of a representative subject at 3T and 7T. On average of 5 subjects, ΔM/M0 values were 0.32±0.05, 0.38±0.06, and 0.19±0.04% at 3T and 0.20±0.02, 0.24±0.02, and 0.11±0.02% at 7T in brain, GM, and WM masks, respectively. ΔM/M0 values were lower at 7T as the result of shorter LD and lower *α*. Based on Eq. 6, *α* at 7T was 70±4% relative to that at 3T. However, SNR increased by 42% from 1.58±0.27 to 2.24±0.28 when comparing 3T and 7T. CBF maps of other subjects can be found in supplement section 12.

**Figure 10.**
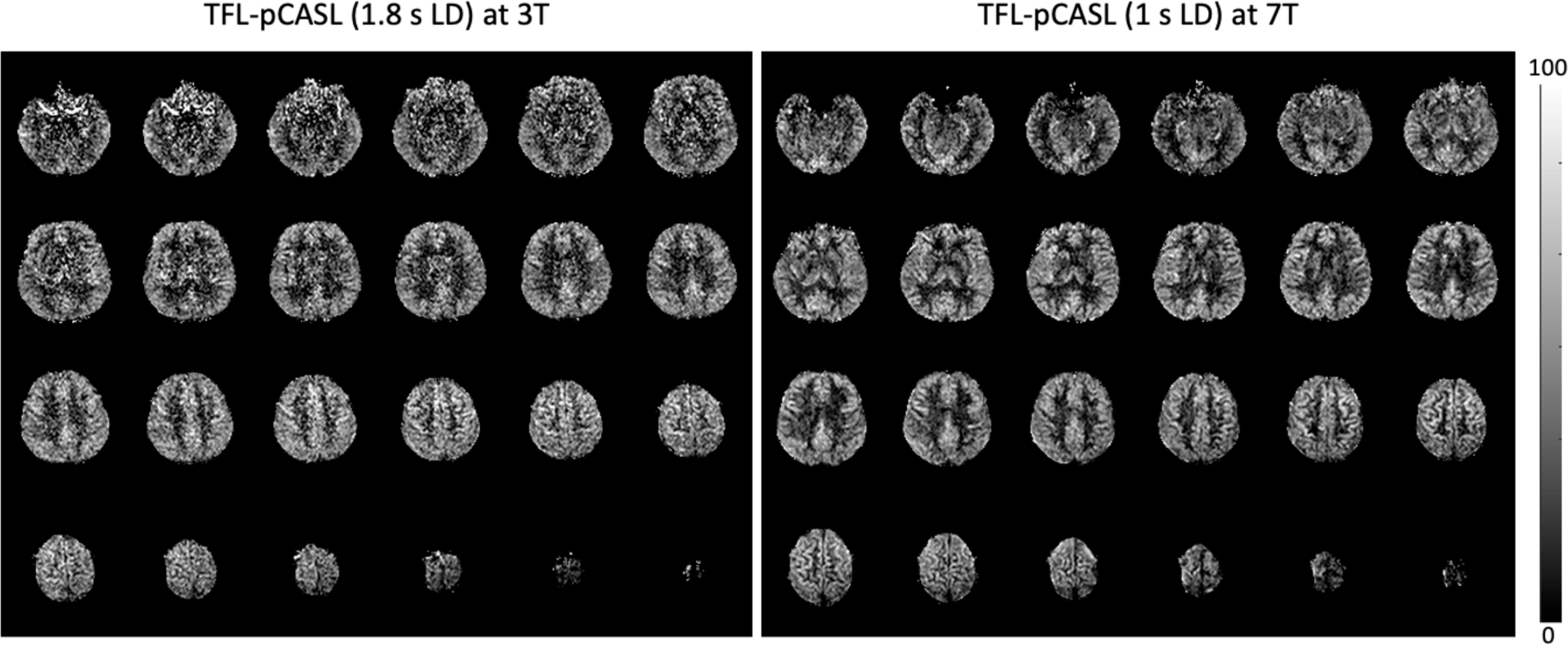
CBF maps (ml/100g/min) acquired with TFL-pCASL at 3T and 7T. While a much longer LD of 1.8 s was used for 3T TFL-pCASL, 7T TFL-pCASL still shows significant improved image quality.

## 4. Discussion

In this study, we presented a distortion-free 3D pCASL sequence with whole-cerebrum coverage at 7T by combining the optimizations of pCASL labeling, BS pulses, and accelerated TFL readout. The proposed pCASL labeling with G_ave_ of 0.4 mT/m and G_ratio_ of 14.67 effectively achieved whole-cerebrum coverage by removing interferences from bottom slices without compromising LE. In addition, optimization tailored for the field inhomogeneities at 7T was conducted for 4 BS pulses, where OPTIM achieved the highest mean IE of 94% and min IE of 92% with a cost of 4.8-fold SAR of the 3T BS pulse. Finally, a 3D TFL readout was presented to acquire pCASL images with 96 mm coverage and 2×2×4 mm^3^ resolution. Compared to GRASE, TFL provided pCASL imaging with comparable SNR and sharpness, no or minimal distortion with fewer artifacts, and easier co-registration with structural MRI. The feasibility for high resolution of 2 mm isotropic pCASL imaging and good to excellent test-retest repeatability of the 2×2×4 mm^3^ protocol were also demonstrated. The proposed TFL-pCASL at 7T also significantly improved SNR when compared to the same sequence at 3T and SMS-TFL-pCASL at 7T.

### 4.1. Optimization of labeling parameters

One of the main contributions of the previous work (Wang et al., 2022) for pCASL labeling was reducing RF spacing and RF duration to minimize the phase accrual between adjacent pulses due to B0 offset. A lower G_ave_ was also suggested to compensate for a reduced B1 field in the labeled neck region at 7T but was unsatisfactory in implementation because of the interference artifacts in bottom slices. Additionally, a single velocity of 40 cm/s was assumed for the labeled blood, which might be oversimplistic. In the current study, we presented a new method to address this artifact and further optimized G_ave_ and G_ratio_ with pulsatile and laminar flow. The simulation yielded Labeling 2 (G_ave_ = 0.4 mT/m and G_ratio_ = 14.67) as the optimal parameter set, effectively eliminating interferences and achieving a comparable LE as the previously proposed Labeling 0 (G_ave_ = 0.6 mT/m and G_ratio_ = 10). In addition, a lower G_ave_ of 0.4 mT/m achieved a higher mean IE both in simulations and in vivo than the original G_ave_ of 0.6 mT/m. To mitigate the influence of B0 offset, compensating gradients during pCASL was also proposed without shortening RF spacing and RF duration (Saïb et al., 2022). However, this method requires additional pre-scans and patient-specific adjustments. It should be noted that B0 correction may be necessary in patients with pathology or metallic implants in the vicinity of the labeling plane

### 4.2. Optimization of background suppression pulses

Using BS is expensive at 7T in terms of SAR and perfusion signal loss. SAR of the two OPTIM pulses constituted 27% of the total SAR of the entire sequence. Although it may not be required for 2D pCASL, BS is necessary for segmented 3D pCASL. As demonstrated in (Alsop et al., 2015), severe artifacts arose as the result of physiological noise and motion during acquisition and between segments. Such artifacts may be more concerning at 7T than at 3T as physiological noise becomes increasingly dominant. Regarding to perfusion signal loss due to BS, we found that, two 3T BS pulse reduced perfusion signal by 80% in the worst case. After optimization, OPTIM significantly reduced the loss to <15% but increased SAR of BS by 4.8-fold. Although the increased SAR led to a longer TR_eff_, SNR efficiency was not compromised given the amount of perfusion signal saved by OPTIM. Alternative methods exist to minimize perfusion signal loss. First, a single BS scheme could be an option to balance the cost and the gain. However, it is hard to achieve satisfactory signal suppression while avoiding negative tissue signals, especially for GM voxels mixing with CSF. Second, a longer LD can compensate for the perfusion signal loss caused by suboptimal BS, for example 1.43 s LD and 3T BS pulse achieve the same SAR as our current protocol. Nevertheless, OPTIM provides uniform IE across whole brain, which is critical for avoiding loss of label due to low IE in inferior brain regions. Moreover, our experiment showed that OPTIM with 1 s LD is already more SNR efficient than 3T BS with 1.43 s LD (supplement section 13). This advantage will be magnified when OPTIM is used with a longer LD. The proposed sequence with OPTIM reached 70%-90% SAR at the first level with a TR of 7 s and a total scan time of 11 min 40 s, which might be acceptable for a clinical scan. For the applications requiring higher temporal resolution, other BS pulses with different performances and SAR levels optimized in this study might be considered based on specific needs.

### 4.3. Evaluation of 3D TFL readout

Compared to GRASE, a drawback of TFL is the longer readout duration. In this study, readout durations were 1344 and 220 ms for TFL and GRASE, respectively. Using a relatively slow TFL sequence for perfusion imaging is feasible for two reasons: 1. TFL can accurately reflect perfusion dynamic at the nominal PLD because k-space center is acquired rapidly in early excitations when centric ordering is used; 2. The assumption that there is no outflow of labeled blood water is valid according to (Alsop et al., 2015; Zhou et al., 2001). Admittedly, a prolonged readout duration may decrease SNR efficiency and cause saturation of labeled blood signal.

However, in our study, a TD (at least 0.9 s) was needed for both 3D TFL-pCASL and GRASE-pCASL to keep SAR levels within the first level. In this case, the SNR efficiency of pCASL was primarily determined by effective TR, which was limited by SAR, instead of the readout duration. TFL-pCASL had a lower SAR than GRASE-pCASL in our in-vivo scans and thus a shorter TR_eff_.

We also applied 2×2 acceleration and segmented readout (N_seg_=2) to reduce TFL readout duration and potential saturation of perfusion signals. Another potential disadvantage of TFL is the intrinsically lower SNR than GRASE because TFL relies on a small FA while GRASE uses FA=90°. However, this SNR difference due to FA is decreased by GRASE’s demand for high acceleration factors and high bandwidth to compensate for T2/T2* relaxation. Also, ASL signal of GRASE-pCASL is more sensitive to the impact of B1 inhomogeneity on refocusing pulses than that of TFL-pCASL (see supplement section 14). Our simulation and in-vivo results indicated 20% (without taking TGV regularization into account) and 40% SNR difference between TFL- and GRASE-pCASL. In addition, there were susceptibility artifacts in GRASE-pCASL, which potentially contributed to lower ICC and wsCV for GRASE-pCASL compared to TFL-pCASL. Although the small sample size (N=5) may lead to underestimated ICC and wsCV, the difference reflected by the measurements was consistent with visual observation of test-retest CBF maps.

Compared to 2D SMS-TFL-pCASL without BS, the proposed 3D TFL-pCASL yielded a higher SNR as a result of thicker excitation volume (20 mm vs 112 mm), constant PLD, centric ordering (1^st^ excitation has 25% higher perfusion signal than 24^th^ excitation), and higher tSNR. Centric ordering was not feasible for SMS-TFL-pCASL due to spiky signals causing interferences between slices. While through-plane blurring does not impact SMS-TFL-pCASL, slice cross-talk had a greater effect on the through-plane image quality of SMS-TFL-pCASL compared to TFL-pCASL. Still, SMS-TFL-pCASL without BS may be a viable approach when SAR is restricted.

Alternative to TFL, integrating balanced steady-state free precession (bSSFP) (Park et al., 2013) and steady-state free precession (FISP) (Gao et al., 2014) with ASL were explored to enable an increased SNR and a minimal saturation of the ASL signal. However, the methods may not be suitable for 7T for two reasons: 1. The banding artifacts could be exacerbated due to B0 inhomogeneity; 2. A low T2/T1 ratio of arterial blood may limit the expected SNR gain.

Compared to TFL-pCASL at 3T, TFL-pCASL at 7T provided 42% higher SNR in similar acquisition time. Given a 0.8 s shorter LD, the improvement achieved by ultrahigh field strength and our optimization is significant.

### 4.4. Limitations and Future Directions

At 7T, parallel transmission (pTx) is an important technique that enables a uniform FA against inhomogeneous B1 distribution with static or dynamic shimming(Padormo et al., 2016). Previous studies tried to improve pCASL labeling, the inversion pulse, and the excitation pulse with pTx (Cloos et al., 2012; Jang et al., 2018; Meixner et al., 2022; Tong et al., 2020; Wang et al., 2022). In this study, we only used the TrueForm method, and the problem of B1 inhomogeneity was tackled by optimizing the adiabaticities of pCASL labeling and BS pulses. As a result, the SNR in the bottom slices remained low because the sinc pulse is prone to a reduced B1. Currently, the use of pTx is limited because it remains difficult to predict local SAR using commercial software. To ensure the safety of subjects, the vendor usually applies conservative estimation of SAR leading to increased scanning time. Future research should take advantage of pTx to fulfill the potential of 7T for ASL.

2D-CAIPIRINHA undersampling and conventional GRAPPA reconstruction were used for the acquisition in this study. More efficient undersampling schemes, such as Poisson-disk (Vasanawala et al., 2011) and stack-of-spirals (Munsch et al., 2020; Vidorreta et al., 2017), and compressed sensing are expected to improve the image quality. Furthermore, the future exploration of the compatibility between TFL and variable flip angle technique (Atkinson et al., 1994) might be valuable for avoiding signal saturation and image blurring. Also, TFL- and GRASE-pCASL were compared using different undersampling pattern and reconstruction methods. Although this may lead to some bias, it is important to compare both sequences with the most optimized imaging parameters and reconstruction method. The two sequences with the same undersampling pattern and GRASE-pCASL with direct inverse FFT reconstruction were presented in supplement section 15. The results were consistent with the finding in section 3.3. Furthermore, no distortion correction was performed for GRASE-pCASL images in this paper. However, as the results of distortion corrected GRASE-pCASL shown in supplement 16, TOPUP of FSL was effective only for correcting geometric distortion, but had no obvious improvement on signal loss and susceptibility artifacts.

Finally, accurate CBF quantification remains challenging at 7T as 𝛼 and physiological parameters, such as arterial transit time, for different groups of people remain to be determined. Future study comparing 7T ASL with 3T ASL and PET are needed to derive these parameters from well-established reference values.

## 5. Conclusion

We presented an optimized pCASL labeling, OPTIM BS pulse, and accelerated 3D TFL readout for 3D pCASL at 7T. The technique yielded a 3D perfusion scan with whole-cerebrum coverage, detailed perfusion and anatomical information without distortion, and adequate image quality. Its clinical value remains to be evaluated in future studies.

## Data Availability

All data produced are available online at http://www.loft-lab.org/index-5.html

## Acknowledgements

This work is supported by National Institute of Health (NIH) grant UF1-NS100614, R01-NS114382, R01-EB032169 and R01-EB028297.

## Data Availability Statement

Experimental data is available upon request at http://www.loft-lab.org/index-5.html. The pulse sequence and reconstruction algorithm described in this work can be requested from the corresponding author through Siemens C2P (Customer to Peer) and University Southern California (USC) Material Transfer Agreement (MTA).

