## Supplement for "Whole-Cerebrum distortion-free three-dimensional pseudo-Continuous Arterial Spin Labeling at 7T"

### Supplementary Information

#### 1. Mathematical expressions of the functions in Section 2.1

##### 1.1 Hanning function:

$$h(t) = \begin{cases} 0.5 + 0.5\cos(2\pi t) & |t| < \frac{1}{2}\tau \\ 0 & \text{otherwise} \end{cases} \qquad \text{Eq.S1}$$

Its Fourier response:

$$H(f) = \tau sinc(\tau f) \cdot \frac{1}{2(1-\tau^2 f^2)}$$

$$f = \gamma G_{max} \cdot z \qquad \text{Eq.S2}$$

$$H(z) = \tau \text{sinc}\,(\tau\gamma G_{max} z) \cdot \frac{1}{2(1-\tau^2(\gamma G_{max} z)^2)}$$

τ is duration of the function, i.e., RF duration, $G_{max}$ is the gradient amplitude during Hanning function, and z is the slice position.

##### 1.2 Temporal sampling function:

For the label condition:

$$x_{la}\,(t) = \sum_{\infty}^{n=-\infty} \delta(t - nT) \qquad \text{Eq.S3}$$

Its Fourier response:

$$X_{la}\,(f) = \sum_{\infty}^{n=-\infty} \delta(f - n\frac{1}{T})$$

$$f = \gamma G_{ave} z \qquad \text{Eq.S4}$$

$$X_{la}\,(f) = \sum_{\infty}^{n=-\infty} \delta(\gamma G_{ave} z - n\frac{1}{T})$$

For the control condition:

$$x_{con}\,(t) = \sum_{\infty}^{n=-\infty} (\delta(t - 2nT) - \delta(t - (2n+1)T)) \qquad \text{Eq.S5}$$

Its Fourier response:

$$X_{con}\,(f) = \sum_{\infty}^{n=-\infty} \delta(f - \frac{1}{2T} - n\frac{1}{T})$$

$$f = \gamma G_{ave} z \qquad \text{Eq.S6}$$

$$X_{con}\,(f) = \sum_{\infty}^{n=-\infty} \delta(\gamma G_{ave} z - \frac{1}{2T} - n\frac{1}{T})$$

T is the duration of a cycle in pCASL labeling, i.e., RF spacing, $G_{ave}$ is the average gradient amplitude in a cycle of pCASL labeling.

##### 1.3 Fourier response of pCASL labeling:

$$H(z) = \hat{f}(h(t) \circledast x(t)) = H(z)X(z) \qquad \text{Eq.S7}$$

$\hat{f}$ represents Fourier transform.

#### 2. The subject cohort for B1 and B0 maps in the imaging volume

This cohort included nine healthy subjects, and the $B_1^+/B_0$ maps of the pCASL imaging volume were acquired with vendor adjustment scans and exported from system files (AdjDataUser.mat and SysDataUser.mat). B1(CP mode) and B0 simming volumes were similar to the pCASL scans. Figure S1 (a) shows the B0 and B1 histograms of this cohort, which supports our selection of optimization ranges for BS (B1 scale: 0.35-1.15 and B0 offset: ±250 Hz). Figure S1 (b) shows the B0 and B1 maps of the pCASL imaging volume from a representative subject.

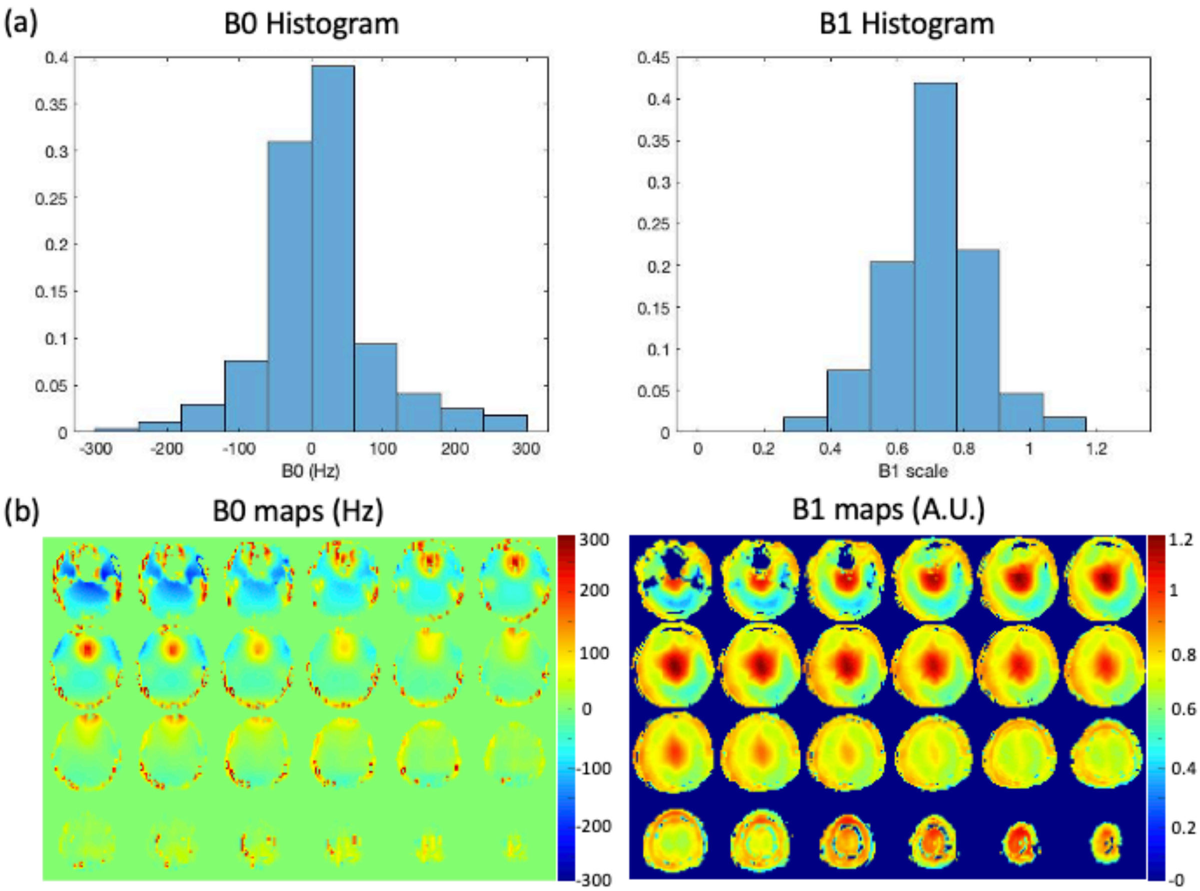


***Figure S1.*** *(a) B0 and B1 histogram from the cohort of nine subjects. (b) B0 and B1 maps from a representative subject.*

#### 3. Optimized BS Pulse definition and parameters

HS pulse

$$|B_1(t)| = Sech(\beta t)$$
$$\Delta\omega(t) = -\mu\beta\tanh(\beta t) \qquad \text{Eq.S8}$$

WURST pulse

$$|B_1(t)| = 1 - |\sin(\frac{\pi}{2}t)|^n$$
$$\Delta\omega(t) = kt \qquad \text{Eq.S9}$$

MATPULSE has a fixed shape that was defined within the software [1]. Only duration and amplitude of the pulse were adjusted in the optimization.

***Table S1.*** *BS pulse parameters*

| | **DURATION (MS)** | **PEAK AMPLITUDE(μT)** | **PULSE-SPECIFIC** | **RELATIVE SAR** |
|---|---|---|---|---|
| **3T PULSE (MATPULSE)** | 10.24 | 10 | / | 1 |
| **OPTIM** | 6 | 20 | / | 4.7 |
| **HS** | 12 | 20 | μ=600, $\beta$=4.4 | 2.3 |
| **WURST** | 11 | 20 | n=2.5, k=650 | 3.7 |
| **MATPULSE** | 9 | 20 | / | 3.1 |

### 4. Theoretical formulization of OPTIM

The RF pulse OPTIM is designed by means of ensemble-based time optimal control [2]. Key component is definition of a robust cost functional for the RF $B_1^+(t) = r(t) \cdot e^{i\varphi(t)}$ with $r(t) = abs(B_1(t))$ and $\varphi = angle(B_1(t))$ as

$$\min_{r,\varphi,T_p} J = T_p + \frac{\alpha}{2}\sum_{i=0}^{N_u} r_i^2 + \frac{\alpha}{2}\sum_{i=0}^{N_u} \varphi_i^2 + \frac{\beta}{p}\sum_{i=1}^{N_{B_1}} \sum_{j=1}^{N_{\Delta B_0}} \left(\frac{M_{i,j}(T_p)-(-1)}{\varepsilon}\right)^p \qquad \text{Eq.S10}$$

with constraint

$$\begin{cases} \frac{dM_{i,j}(t)}{dt} = A_{i,j} \cdot M_{i,j}(t) + b, \\ M_{i,j}(0) = (0,0,1)^T, \forall i = 1,\cdots N_{B_1}, \forall j = 1,\cdots N_{\Delta B_0}, \forall t \in [0, T_p], \\ 0 \le r(t) \le r_{\max}, 0 \le \varphi(t) < 2\pi. \end{cases} \qquad \text{Eq.S11}$$

Therein, $T_p$ is the pulse duration to be minimized, and α (>0) is the regularization parameter. The last term within Eq.S10 ensures inversion efficiency of the z-component of the magnetization $M_{i,j}$ (penalization parameter β>0). Hereby, $N_{B1}$ discrete values of $B_1$+ scales are assumed, and $N_{\Delta B_0}$ different instances of $B_0$ offsets. The parameter $\varepsilon$ defines the gap to the desired magnetization, i.e. determines quality of the inversion within an $L^p$-norm, p≥2, p is even. The Bloch equations Eq.S11 are solved for all instances i of $B_1$+ scales, and offsets j of B0 by symmetric operator splitting [3]. Within the last row of Eq.S11, box constraints on the RF magnitude and phase are defined.

The optimization itself is performed within a trust-region, semi-smooth quasi-Newton method based on exact discrete derivatives supplied by adjoint calculus [4].

#### 5. Labeling plane and B0 shimming

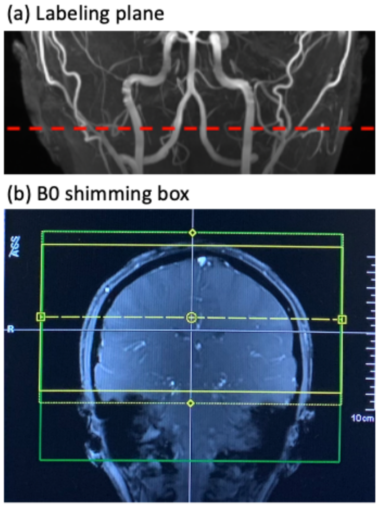


***Figure S2.*** *(a) Labeling plane was placed at the C1 segment (Bouthillier classification) of the internal carotid arteries. (b) B0 shimming box (green) covered the whole imaging volume with an extended bottom edge to cover labeling plane.*

#### 6. Calculation of IE map

Single slice 2D TFL sequences with and without a BS pulse were used to obtain IE maps of the BS pulses. The sequence with a readout immediately following one of BS pulses was employed to acquire the first image (S1). Then, a control sequence without the BS pulse was used to acquire the second image (S2). $TR_{eff}$ was 10 s for both sequences to allow a full recovery of magnetization. The two images had the same FOV covering whole brain. An IE map can be obtained as S1/S0 with some additional scaling and intercept, which can be calculated as follows,

$$\boldsymbol{S_{1/2}} = \left|\left[\boldsymbol{M_{1/2}}(\boldsymbol{E_1}\cos\boldsymbol{\theta})^{j-1} + (1-\boldsymbol{E_1})\frac{1-(\boldsymbol{E_1}\cos\boldsymbol{\theta})^{j-1}}{1-\boldsymbol{E_1}\cos\boldsymbol{\theta}}\right]\boldsymbol{E_2}\sin\boldsymbol{\theta}\,\right| \quad \text{Eq. S12}$$

Let K = $(\boldsymbol{E_1}\cos\ \boldsymbol{\theta})^{j-1}$ and P = $(1-\boldsymbol{E_1})\frac{1-(\boldsymbol{E_1}\cos\ \boldsymbol{\theta})^{j-1}}{1-\boldsymbol{E_1}\cos\ \boldsymbol{\theta}}$, and assume $\boldsymbol{M_2} = 1$ and $\boldsymbol{M_1}$ is negative

$$\frac{S_1}{S_2} = \frac{M_1K+P}{K+P} \quad \text{Eq. S14}$$

$$IE = -M1 = \frac{S_1}{S_2}\frac{K+P}{K} - \frac{P}{k} \quad \text{Eq. S15}$$

where Eq. S12 is based on [5], $\boldsymbol{E_1} = e^{-\frac{ES}{T1}}$, $\boldsymbol{E_2} = e^{-\frac{TE}{T2}}$, T1 was set to 1500 ms, T2 was set to 50 ms, j was set to 16 for linear ordering, and FA $\boldsymbol{\theta}$ was 8°. The experiment was repeated for axial, sagittal, and coronal views to evaluate all BS pulses.

### 7. Simulated PSFs in PE direction

There was no significant difference of FWHM in PE direction between the two methods. FWHM values were 1.06 and 1.08 for TFL-pCASL and GRASE-pCASL, respectively.

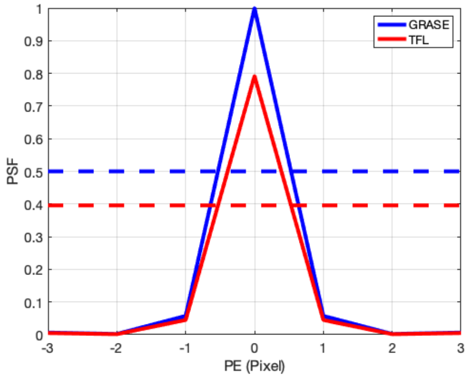


***Figure S3.*** *Simulated PSFs of GRASE and TFL in PE direction.*

### 8. Coregistration of TFL- and GRASE-pCASL on T1

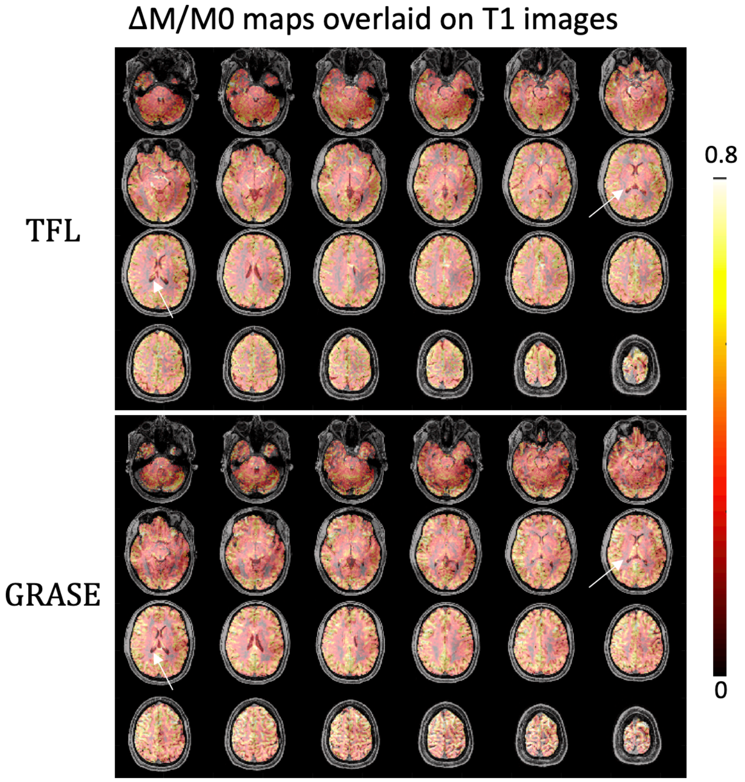


***Figure S4.*** *T1 images were coregistered to the M0 images of TFL-pCASL and GRASE-pCASL using SPM12, and ΔM/M0 maps overlaid on T1 images were shown. ΔM/M0 maps of TFL-*

*pCASL matched perfectly with T1 images, whereas that of GRASE-pCASL showed some distortions in bottom slices and frontal lobes. In addition, some detailed anatomical information was less accurate in GRASE-pCASL compared to TFL-pCASL. As indicated by white arrows, there was a slight shift of the hippocampus between GRASE-pCASL and T1 images.*

### 9. Higher resolution (2 mm isotropic) protocol

A 2 mm isotropic high-resolution protocol was tested with adjusted parameters, including resolution = 2×2×2 mm$^3$, FOV = 224×192×104 mm$^3$, matrix size = 112×96×52, 4 oversampled slices, $N_{seg}$ = 4, 26 measurements, including an M0 image, acquired in 12 min 10 s. Figure S5 (a) and (b) display ΔM/M0 maps with a standard 2×2×4 mm resolution and a 2 mm isotropic high resolution, respectively. In Fig. S5 (c), higher resolution revealed finer through-plane details and more accurate delineation of GM/WM boundaries than standard resolution. However, SNR was lower with the finer resolution.

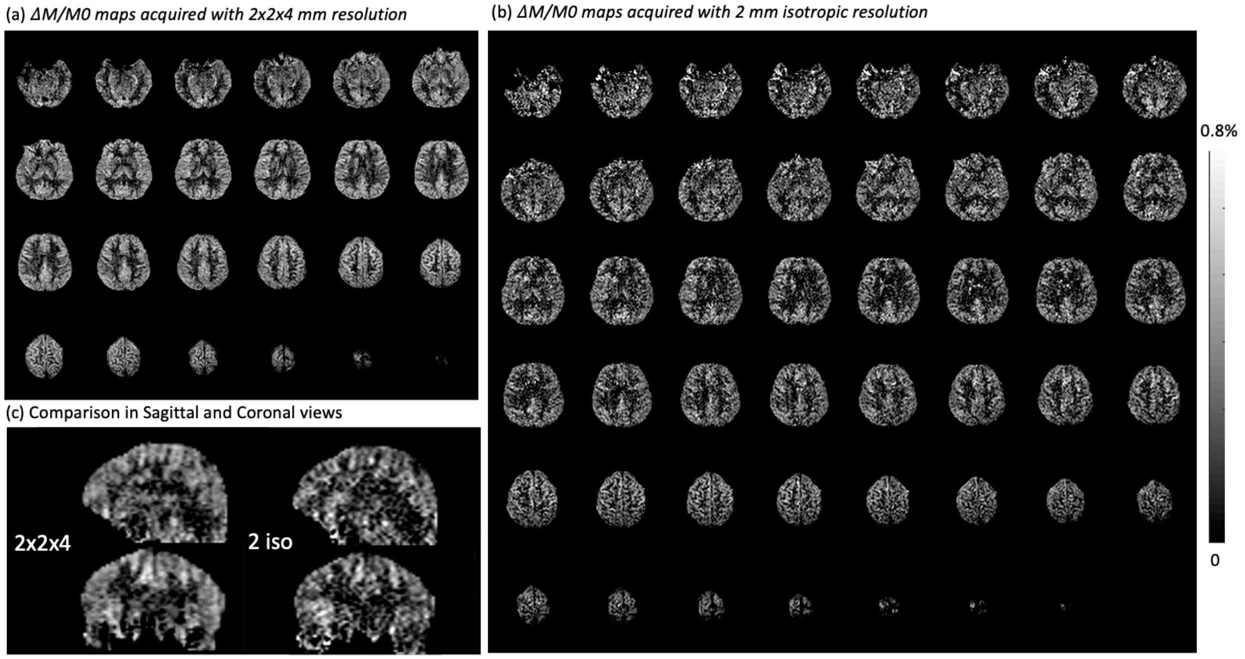


***Figure S5.*** *(a) and (b) shows* ***Δ****M/M0 maps acquired with 2×2×4 mm3 and 2 mm isotropic resolutions on the same subject. Anatomical details, such as orbitofrontal cortex, choroid plexus, cerebellum, and gyri, can be discerned. (c) Comparison of the two scans in Sagittal and Coronal views. Finer through-plane details can be observed with 2 mm isotropic resolution.*

### 10. Test-retest repeatability

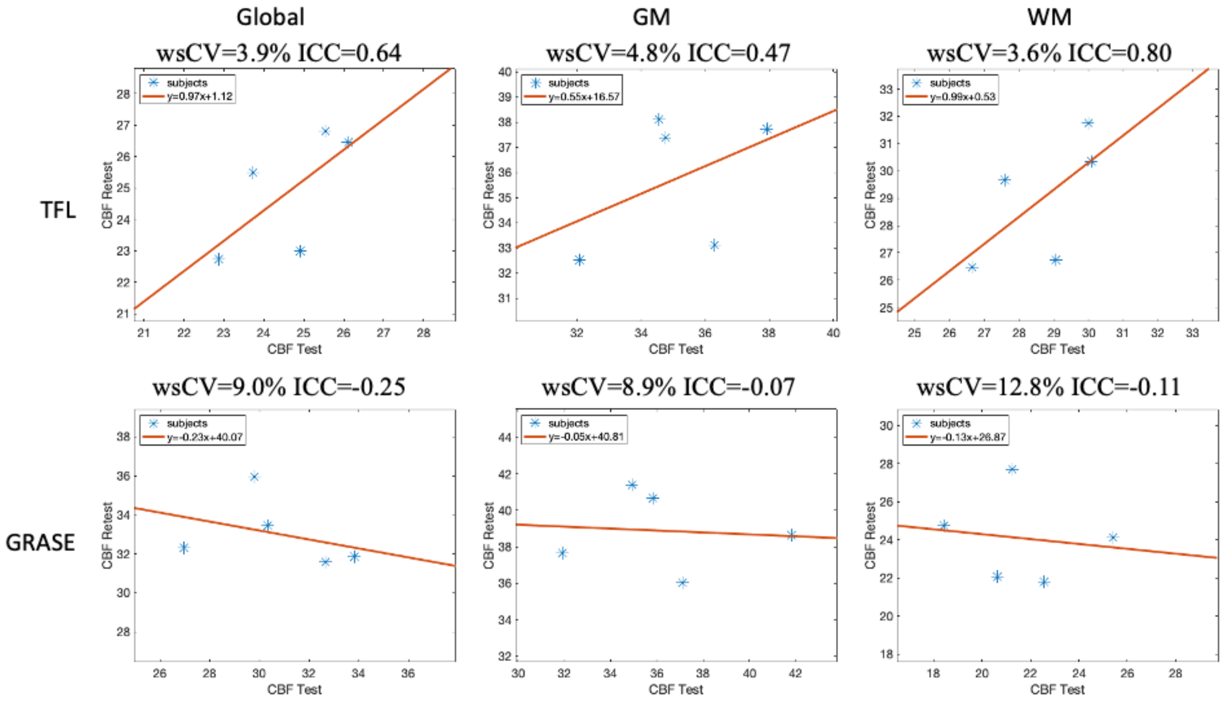


***Figure S6***. *Scatter plots of test-retest results of TFL- and GRASE-pCASL. wsCV and ICC were listed.*

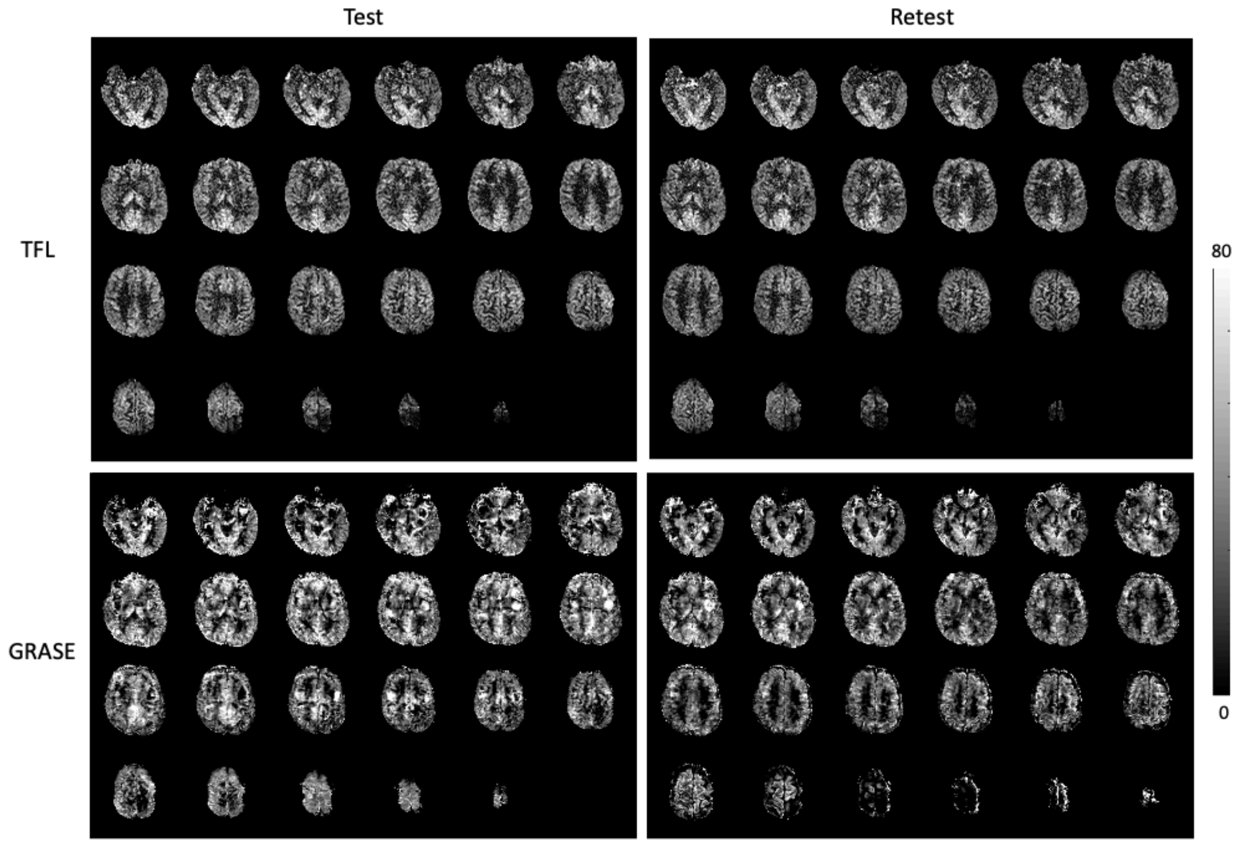


***Figure S7****. CBF (ml/100g/min) maps of subject 2 in two visits using* TFL- and GRASE-pCASL.

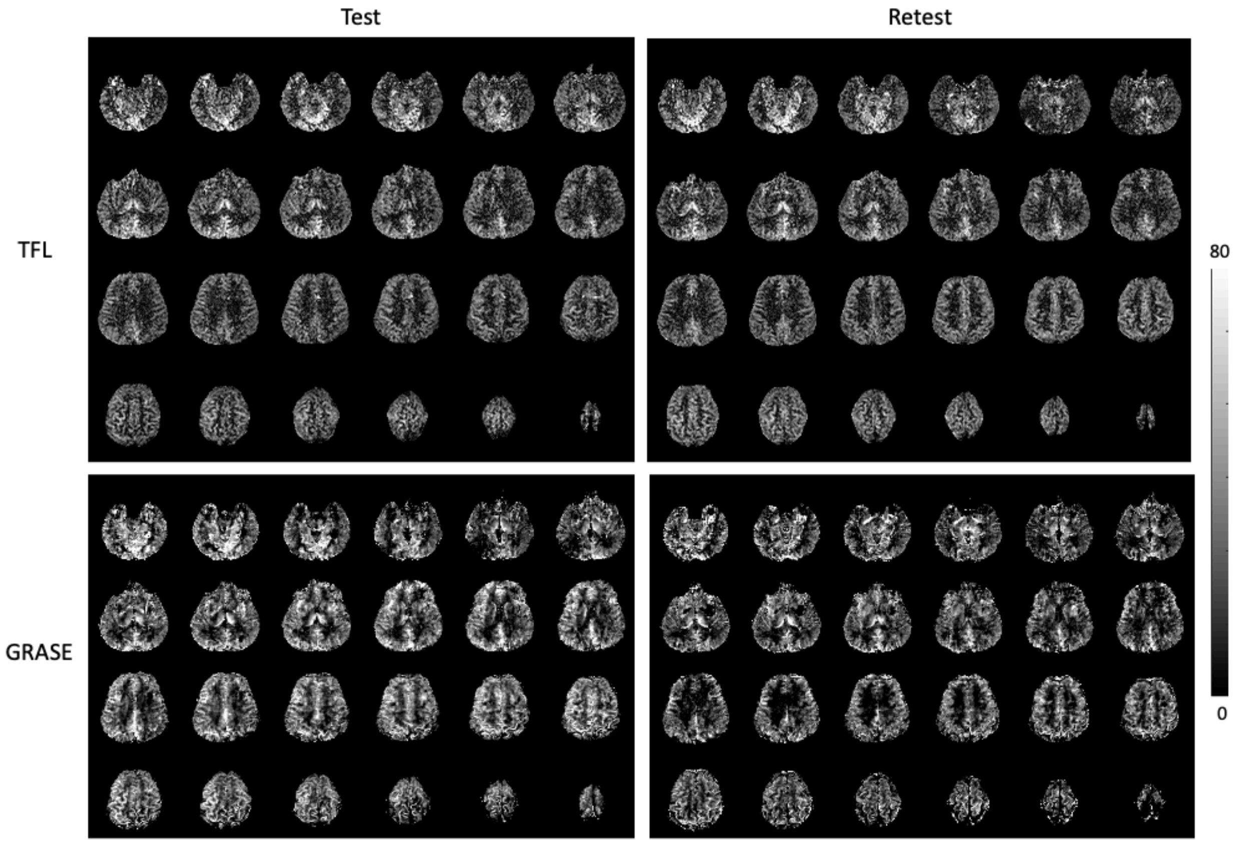


***Figure S8***. *CBF (ml/100g/min) maps of subject 3 in two visits using* TFL- and GRASE-pCASL.

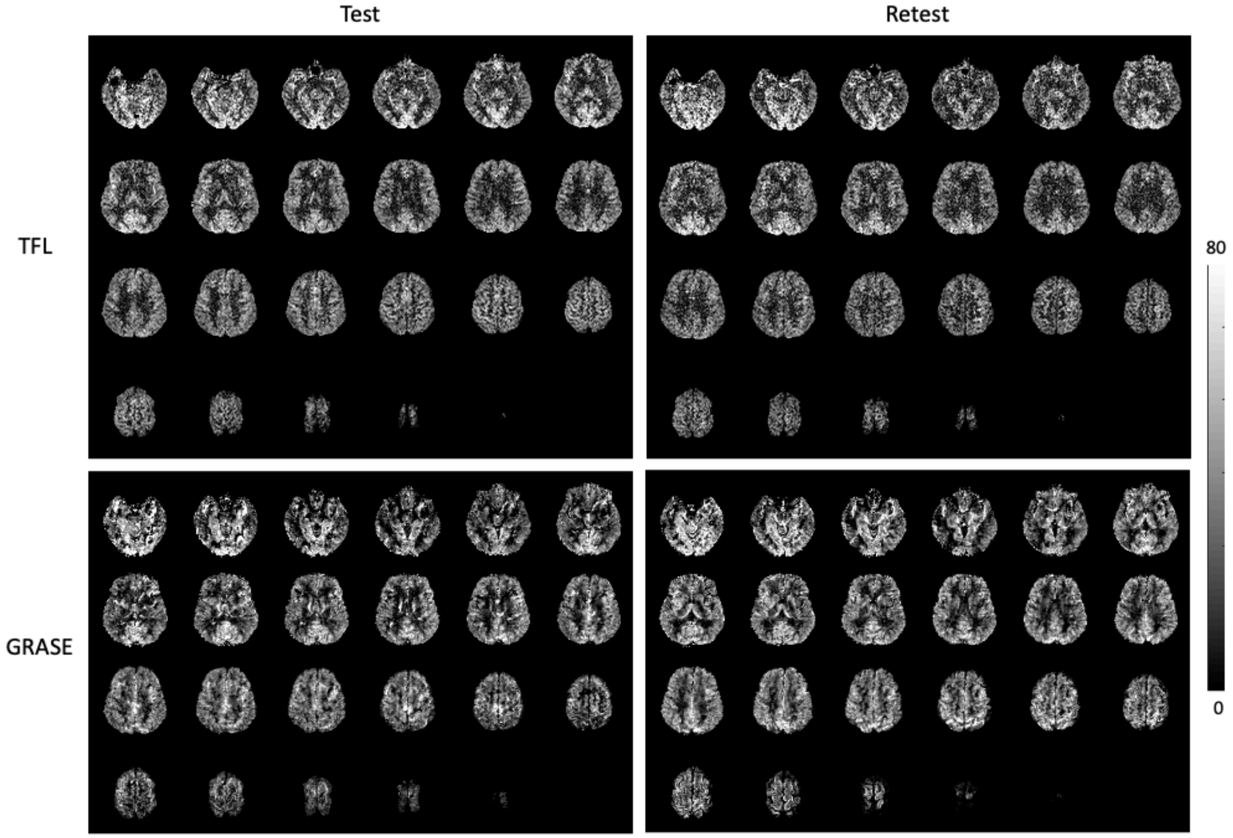


***Figure S9***. *CBF (ml/100g/min) maps of subject 4 in two visits using* TFL- and GRASE-pCASL.

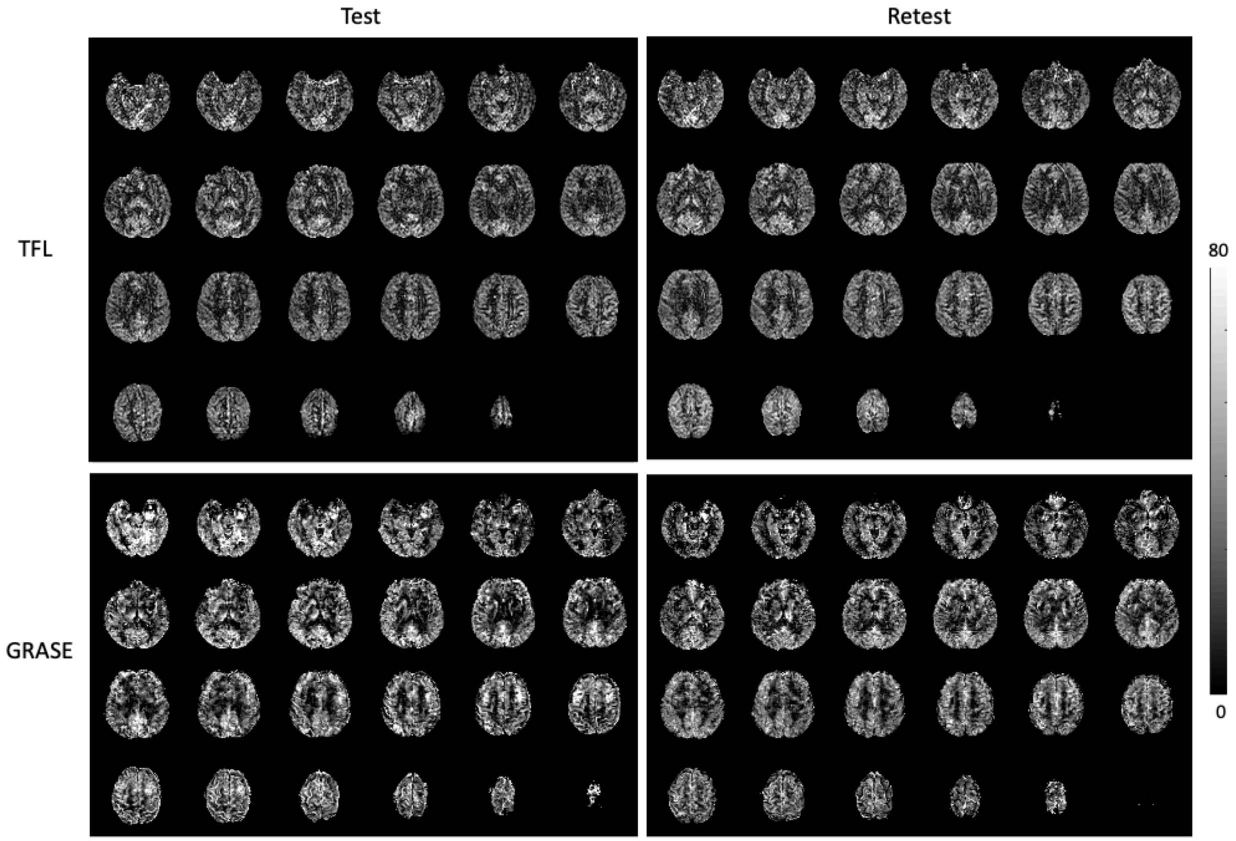


***Figure S10***. *CBF (ml/100g/min) maps of subject 5 in two visits using* TFL- and GRASE-pCASL.

### 11. CBF and SD maps of 2D SMS-pCASL

CBF can be quantified for 2D SMS-pCASL based on Eq. 6 taking different PLDs (245 ms delay between slice groups) for the 5 slice groups. In addition, j in Eq. 6 was set to 24 because k-space center was acquired at the $24^{th}$ excitation. Figure S11 compares CBF maps of a subject acquired by 2D SMS-TFL-pCASL and 3D TFL-pCASL. Although intensity decay can be corrected for later slice groups of SMS-TFL-pCASL, nosier appearance can be still observed.

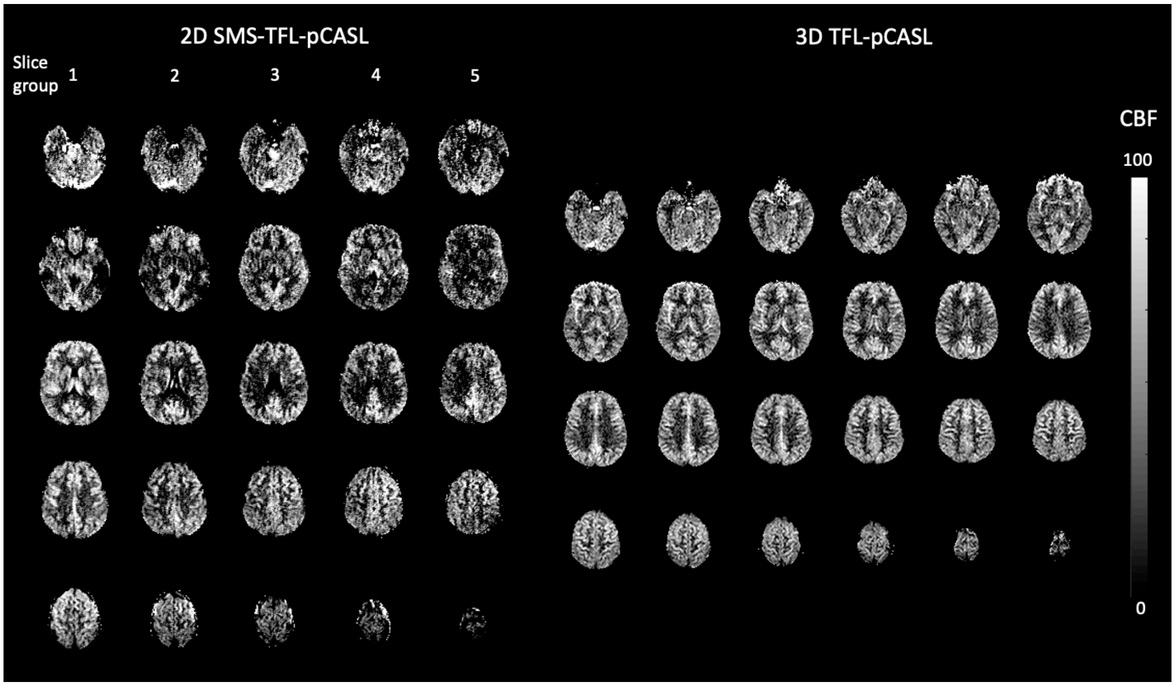


***Figure S11****. CBF (ml/100g/min) maps of 2D SMS-TFL-pCASL and 3D TFL-pCASL.*

SNR of SMS-TFL-pCASL may also be further affected by reduced tSNR due to the lack of BS. tSNR values in the brain mask were 0.31 and 1.07 for SMS-TFL-pCASL and 3D TFL-pCASL, respectively. Figure S12 compares the temporal noise SD maps of perfusion images acquired with SMS-TFL-pCASL and 3D TFL-pCASL. Due to substantially higher intensity of the SD map of SMS-TFL-pCASL than its PWI, the intensity of two SD maps were scaled down by a factor of 10. Figure S12 shows overwhelming temporal noise from CSF and tissues for SMS-TFL-pCASL, while only some inflow vessel signal can be seen in 3D TFL-pCASL. The result may suggest that BS is also necessary for SMS-TFL-pCASL.

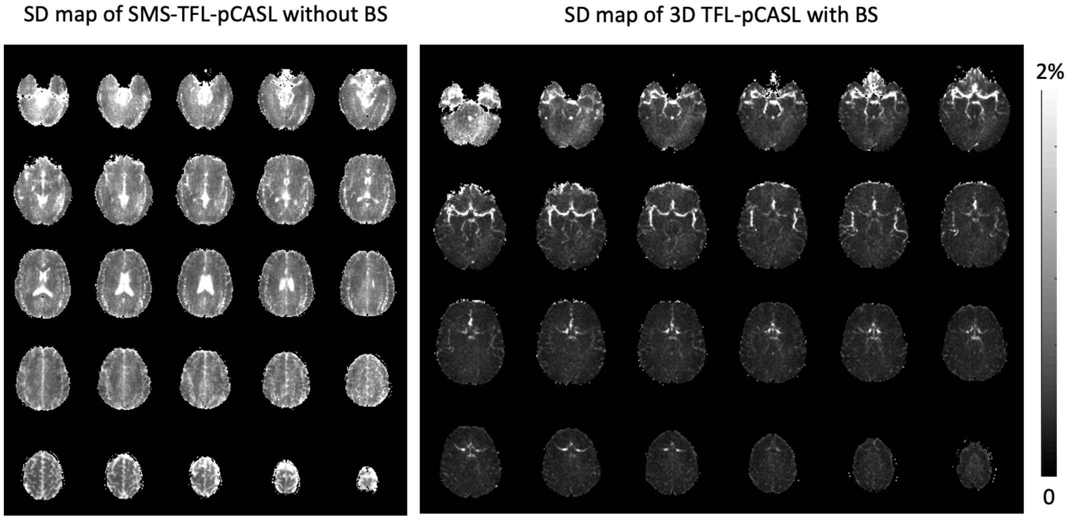

***Figure S12.*** *Temporal noise SD maps calculated from a series of individual ΔM/M0 maps acquired by 2D SMS-TFL-pCASL without BS and 3D TFL-pCASL with BS.*

### 12. Comparison of 3T and 7T

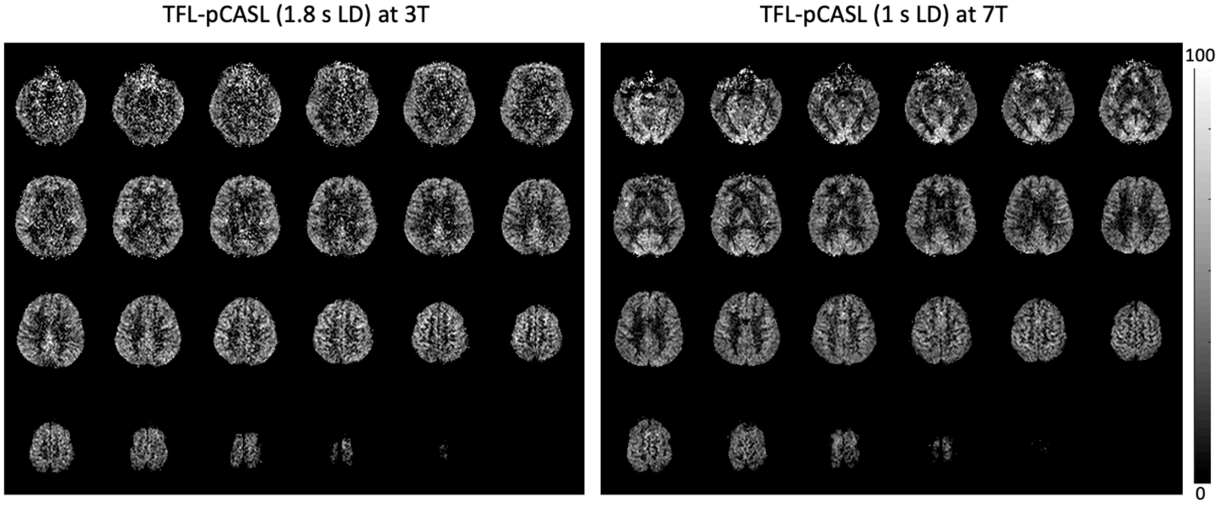


***Figure S13***. *CBF (ml/100g/min) maps of subject 2 acquired by TFL-pCASL at 3T and 7T, respectively. 3T ASL used 1.8 sec LD, while 7T ASL used 1 sec LD.*

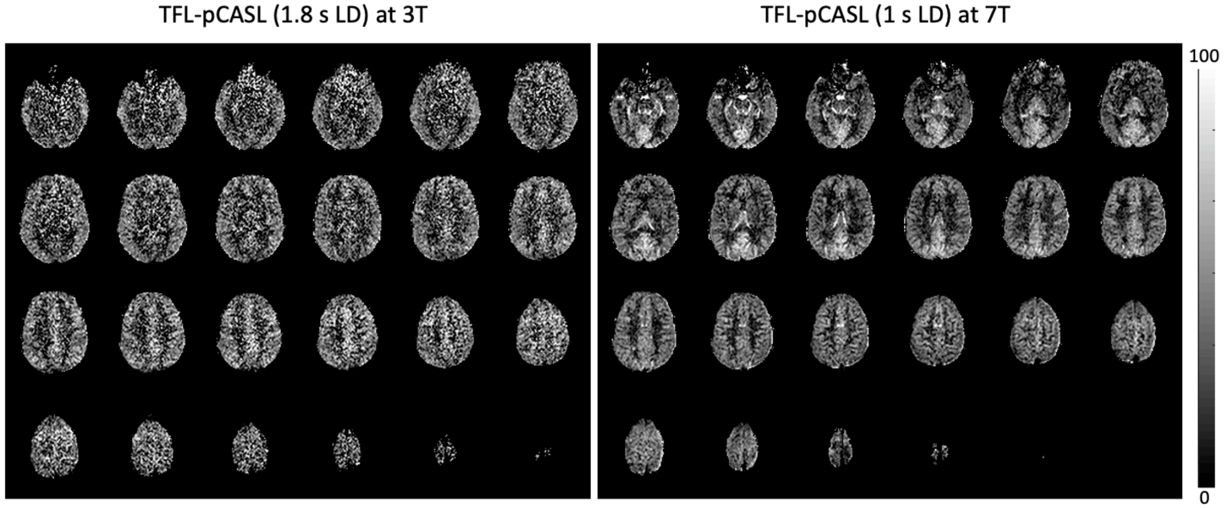


***Figure S14***. *CBF (ml/100g/min) maps of subject 3 acquired by TFL-pCASL at 3T and 7T, respectively. 3T ASL used 1.8 sec LD, while 7T ASL used 1 sec LD.*

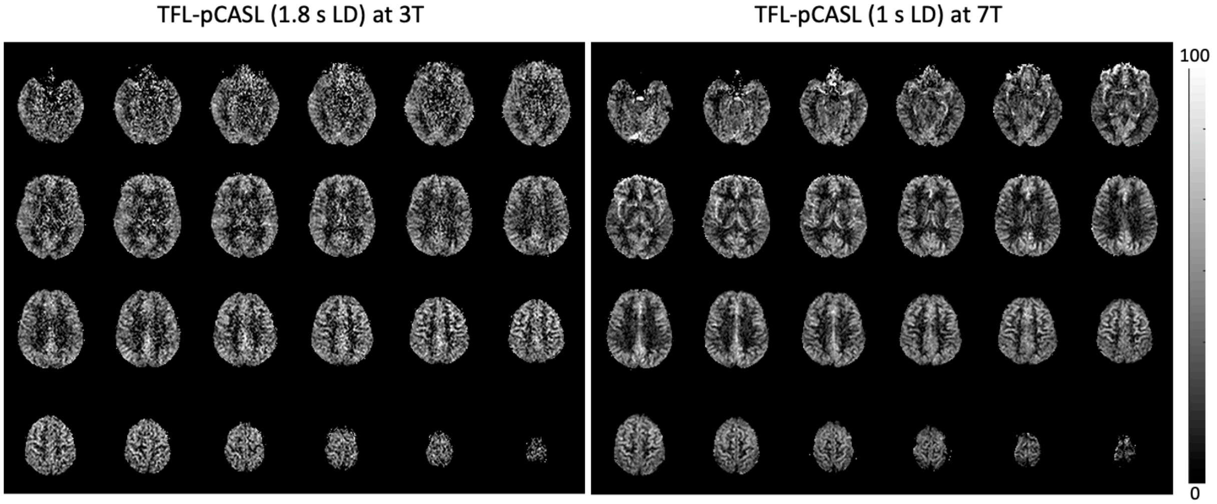


*Figure S15*. CBF (ml/100g/min) maps of subject 4 acquired by TFL-pCASL at 3T and 7T, respectively. 3T ASL used 1.8 sec LD, while 7T ASL used 1 sec LD.

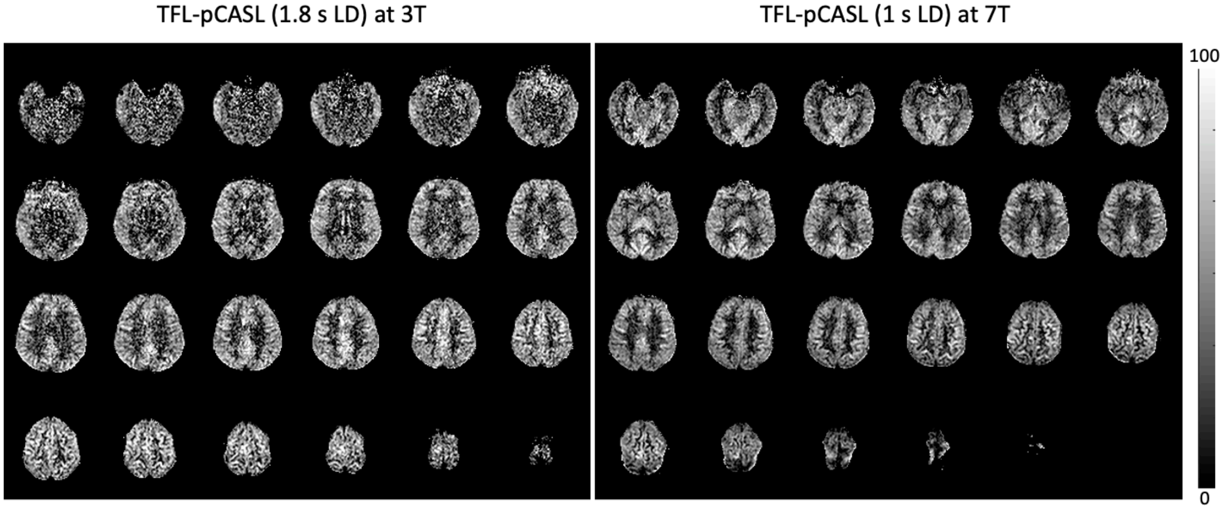


*Figure S16*. CBF (ml/100g/min) maps of subject 4 acquired by TFL-pCASL at 3T and 7T, respectively. 3T ASL used 1.8 sec LD, while 7T ASL used 1 sec LD.

### 13. Comparison of the default protocol and a protocol using 3T BS and 1430 ms LD

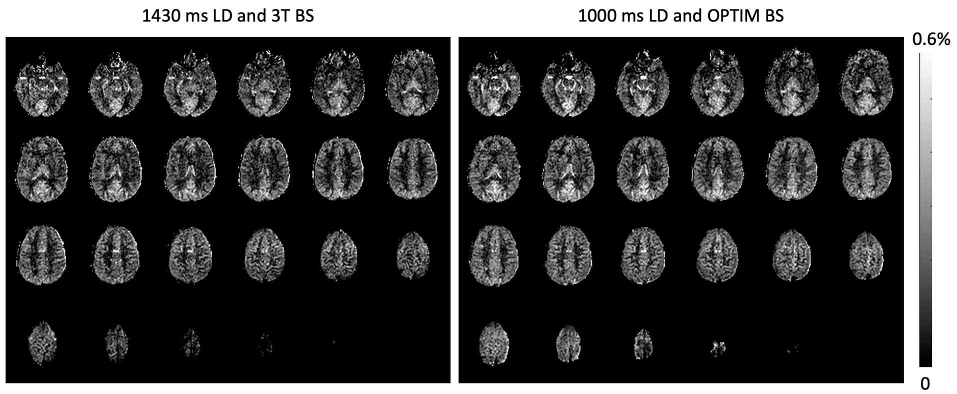


***Figure S17.*** *ΔM/M0 maps acquired with two protocols that used 1430 ms LD and 3T BS and 1000 ms LD and OPTIM BS, respectively. The two protocols had the same SAR. The measured ΔM/M0 values in the GM mask were 0.19% and 0.21% for the former and the latter protocol, respectively.*

### 14. Simulation of ASL signal of TFL- and GRASE-pCASL under B1 inhomogeneity

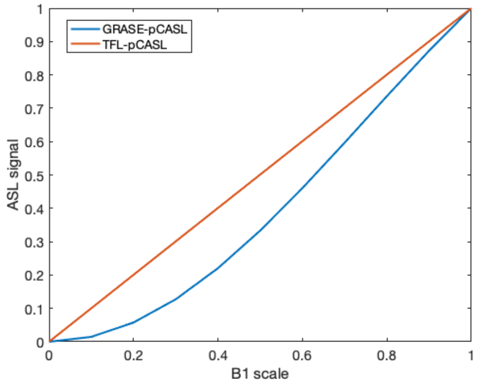


***Figure S18****. Relationships between ASL signal and B1 scale for TFL- and GRASE-pCASL that were simulated based on Eq. 4 and 5, respectively. For each sequence, signal intensities were normalized by the signal intensity that was obtained with the nominal FA (B1 scale = 1).*

### 15. GRASE-pCASL using same sampling pattern as TFL-pCASL

In our comparison study, TFL- and GRASE-pCASL used different sampling patterns and reconstruction methods. We believe it is important to compare TFL-pCASL with the best available GRASE-pCASL option. The reason that we chose the single segment TGV reconstruction for GRASE-pCASL is that this setup is robust to physiological fluctuation between segments and is more SNR efficient than segmented TGV reconstruction and full-sampled inverse FFT reconstruction according to [6]. Also, the reason that we chose the undersampling pattern with R=3x3 instead of R=4x2 (identical to TFL-pCASL if two control/label pairs were combined as segments) for GRASE-pCASL is that R=3x3 significantly decreases the number of spin echoes from 14 to 10 with the cost of 4 ms longer TE compared to R=4x2.

Nevertheless, here we present the GRASE-pCASL CBF maps acquired with the R=4x2 undersampling and reconstructed with three reconstruction options, including single segment TGV reconstruction, two segments TGV reconstruction, and eight segments IFFT reconstruction. Imaging parameters were: resolution = 2.1×2.1×4 $mm^3$, FOV = 200×200×112 $mm^3$, matrix size = 112×92×28, 6 oversampled slices, bandwidth = 2170 Hz/pixel, echo spacing = 0.53 ms, TE = 18 ms, 2D-CAIPIRINHA with R=4×2, EPI factor = 23, turbo factor = 14, single shot, FA=120$^{\circ}$, TR = 8 s, 88 measurements, including an M0 image, were acquired in 11 min 40 s. Distortions in GRASE images were corrected by using a blip-reversed scan and TOPUP (details see supplement section 15).

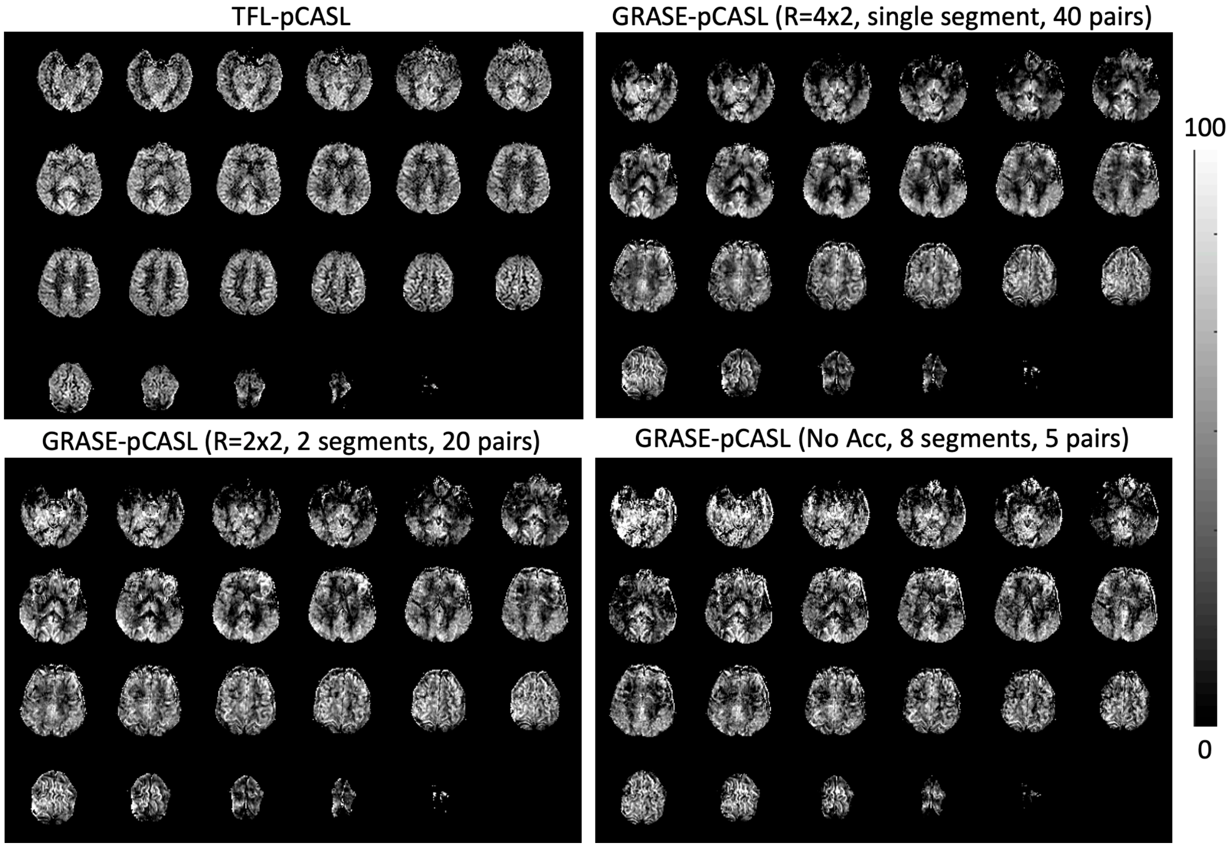


***Figure S19***. *CBF maps for a comparison of TFL-pCASL and GRASE-pCASL with R=4x2 and three different reconstruction options, including single segments, two segments, and 8 segments.*

Figure S19 shows the comparison of the CBF maps acquired by TFL-pCASL and GRASE-pCASL with R=4x2 and three different reconstruction options. Compared to TFL-pCASL, all GRASE-pCASL images showed artifacts and signal dropouts caused by susceptibility effects. Although similar image appearance was observed among GRASE-pCASL images reconstructed by three reconstruction options, the single segment image with TGV reconstruction showed the highest SNR. The global SNR values were measured as 2.41 for TFL-pCASL, and 3.82, 2.29, and 1.14 for GRASE-pCASL with 1, 2, and 8 segments, respectively.

### 16. Distortion correction for GRASE-pCASL

Distortion correction was attempted to improve the image quality of GRASE-pCASL that was described in supplement section 14 (R=4x2, single segment). A separate blip-reversed scan with PE direction of posterior to anterior (PA) was acquired, while the PE direction of GRASE-pCASL scans was AP. The TOPUP function in FSL [7] was used to correct distortion with the proper input parameters, such as blip directions and total readout time of 12.2 ms (defined as the center of first EPI echo to the center of the last EPI echo).

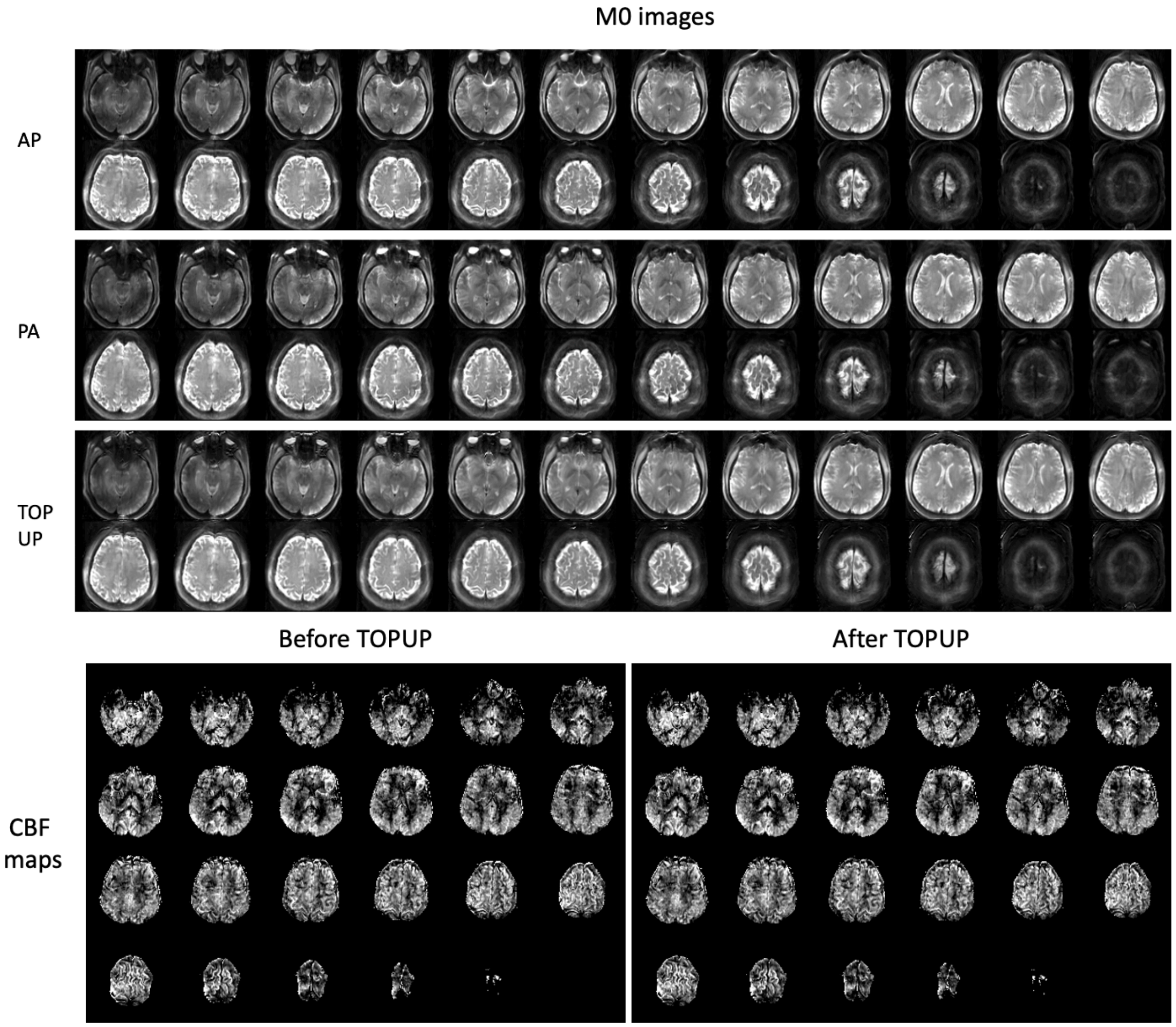


***Figure S20***. *Blip-reversed M0 images and the M0 images after distortion correction, and CBF maps before and after TOPUP.*

Figure S20 shows blip-reversed M0 images and the M0 images after distortion correction, and CBF maps before and after TOPUP. Although TOPUP successfully corrected distortions, the signal loss related to susceptibility effects was not restored, which was reflected in the CBF maps. There was no apparent difference between the CBF maps before and after TOPUP.
